# Transmission of SARS-CoV-2 from pre and asymptomatic infected individuals. A systematic review

**DOI:** 10.1101/2021.07.28.21261254

**Authors:** Tom Jefferson, Elizabeth A. Spencer, Jon Brassey, Igho J. Onakpoya, Elena C. Rosca, Annette Plüddemann, David H. Evans, John M. Conly, Carl J. Heneghan

**Affiliations:** Department for Continuing Education, University of Oxford, Rewley House, 1 Wellington Square, Oxford OX1 2JA, UK; Centre for Evidence Based Medicine, Nuffield Department of Primary Care Health Sciences, University of Oxford, Radcliffe Observatory Quarter, Oxford, OX2 6GG, UK; Trip Database Ltd, Glasllwch Lane, Newport, NP20 3PS, UK; Victor Babes University of Medicine and Pharmacy, Piața Eftimie Murgu 2, Timișoara 300041, Romania; Li Ka Shing Institute of Virology and Dept. of Medical Microbiology & Immunology, University of Alberta; Departments of Medicine, Microbiology, Immunology & Infectious Diseases, and Pathology & Laboratory Medicine, Synder Institute for Chronic Diseases and O’Brien Institute for Public Health, Cumming School of Medicine, University of Calgary and Alberta Health Services, Calgary, Canada.

**Keywords:** SARS-CoV-2, Transmission, Levels of evidence, Asymptomatic cases, Presymptomatic cases.

## Abstract

**Background:** The transmission role of SARS-Cov-2 infected persons who develop symptoms post testing (pre symptomatics) or not at all throughout the course of positivity (asymptomatics) is unknown. We carried out a systematic review of available evidence to determine whether they were infectious or not and if so for how long and their probable contribution to the pandemic spread of SARS-CoV-2.

**Methods:** We searched LitCovid, medRxiv, Google Scholar and the WHO Covid-19 databases and reference lists of included studies. Search terms were COVID-19, SARS-CoV-2, transmission, asymptomatic, presymptomatic and appropriate synonyms. Searches were carried out to 31 March 2021. We included studies on people exposed to SARS CoV-2 within 2-14 days (incubation time) of close contact or suspected community or institutional exposure to index asymptomatic (at the time of observation) infected individuals, as defined in the study. We included studies with a proven or hypothesised chain of transmission with secondary case infected based on fulfilling a confirmed or probable case definition and confirmation of infectiousness and transmission outcome based either on serial PCR cycle threshold readings or viral culture or gene sequencing or any combination thereof and adequate follow up. We assessed the reliability of eliciting symptom and signs compatible with contemporary knowledge and extracted documentation of the likelihood of transmission, presence of replicating virus and/or documentation of phylodynamics (genetic sequence lineage) and/or adequate follow-up and reporting of symptoms and signs. We wrote to all included studies corresponding authors to request further details and assessed likelihood of transmission using adapted causality criteria.

**Results:** We included 18 studies from a variety of settings. Because of the current lack of standardized methodology and clear reporting criteria there was substantial methodological variation in transmission studies. Asymptomatic prevalence at the time of initial testing varied from 12.5% to 100% and of these 6% to 100% were pre-symptomatic cases, depending on the setting and the methods of case ascertainment and the population. Nursing/care home facilities reported high rates of presymptomatic: 50% - 100% (n=3 studies). Fifteen studies were classified as high risk and three studies at moderate risk of symptom ascertainment bias. In practice, this assessment means that high-risk studies may be less likely to distinguish between pre-symptomatic and asymptomatic cases. Six of the asymptomatic studies and four presymptomatic studies reported growing infectious virus although the data was too sparse to determine duration of infectiousness. Three studies were judged as providing possible and three of probable/likely evidence of asymptomatic transmission of SARs-CoV-2. Five studies provided evidence of possible and two of probable/likely presymptomatic transmission of SARs-CoV-2. Author response rate was 100%.

**Conclusions:** Reliable studies included here provide probable evidence of transmission of SARS-CoV-2 from presymptomatic and asymptomatic individuals. Single point in time estimates and binary PCR testing alone cannot provide reliable information on symptom status and information on infectivity. The number of studies and asymptomatic and presymptomatic cases eligible for inclusion was low, with more data and international standardisation of methods needed to further reduce uncertainty.

## Introduction

The overarching aim of the WHO’s <u>Global Strategic Preparedness and Response Plan for COVID-19</u> is to prevent transmission of SARS-CoV-2 and prevent associated illness and death. However, the transmission of the SARS-CoV-2 virus and the disease it causes are not entirely understood, and public health and social measures (PHSMs) for restricting transmission are based on limited data with relatively few high-quality systematic reviews on the transmission of the SARS-CoV-2 virus available. To date, systematic reviews have revealed methodological shortcomings in the included studies that hinder the development of firm conclusions over the transmission dynamics^1–5^.

Several reviews have addressed the prevalence of asymptomatic COVID-19 cases but the design and reporting of studies included in those reviews reveal multiple deficits and biases, which impact the asymptomatic estimates, highlighting the need for more robust evidence^5, 6^. Limitations identified include relying on binary PCR testing alone to estimate asymptomatic fractions and onward transmission^7, 8^. Previous estimates of the asymptomatic influenza fraction are similarly affected by low quality study designs and methods. In addition, the role of cases that remain without symptoms or signs throughout the active phase (asymptomatic) of illness and those who have not developed symptoms or signs yet when surveyed (pre-symptomatic) is at present unclear, partly because of limitations in the methodologies employed in the studies^5, 9^.

A lack of standardised methods requires integrating clinical, epidemiologic, molecular and laboratory evidence into a framework that identifies higher-quality evidence to reduce the uncertainty over the transmission dynamics of acute respiratory pathogens, including SARS-CoV-2. The framework requires studies that use comprehensive and serial screening for symptoms^5, 9 10^ and the use of high-level confirmatory evidence of infection including viral culture and/or whole-genome sequencing to indicate replicable, infectious virus or confirmation of identical sequences and a robust epidemiologic link^10^. The framework is a work in progress as scientific understanding in this area evolves.

### Objectives

To provide a rapid summary and evaluation of relevant data on the transmission of SARS-CoV-2 from pre and asymptomatic individuals, report essential policy implications, and highlight research gaps that require attention.

## Methods

This review is part of a series of living reviews^1–4^ updated as new and vital research is published. The review protocol is available at: medRxiv 2021.05.06.21256615; doi: https://doi.org/10.1101/2021.05.06.21256615. We set out to address the following questions:

1. Are asymptomatic or presymptomatic PCR positive individuals infectious;
2. If asymptomatic PCR positive individuals are infectious, what proportion are infectious and what is the duration of infectiousness;
3. What is the relationship between infectiousness and PCR cycle threshold;
4. Is there evidence of a chain of transmission that establishes asymptomatic and/or presymptomatic transmission of SARs-CoV-2?

## Search Strategy

The following electronic databases were searched: LitCovid, medRxiv, Google Scholar and the WHO Covid-19 database. Search terms were COVID-19, SARS-CoV-2, transmission, asymptomatic, presymptomatic and appropriate synonyms. Also, the reference lists of included studies were searched for additional relevant studies. Searches were carried out up to 31 March 2021.

### WHO Covid-19 Database

(https://search.bvsalud.org/global-literature-on-novel-coronavirus-2019-ncov/). The global literature cited in the WHO COVID-19 database is updated daily (Monday through Friday) from searches of bibliographic databases, hand searching, and the addition of other expert-referred scientific articles.

### LitCovid

(https://www.ncbi.nlm.nih.gov/research/coronavirus/)

A curated literature hub for tracking up-to-date scientific information about the 2019 novel Coronavirus. It is a comprehensive resource on the subject, providing central access to relevant articles in PubMed.

### MedRxiv

(https://www.medrxiv.org/)

A free online archive and distribution server for complete but unpublished manuscripts (preprints) in the medical, clinical, and related health sciences.

### Google Scholar

(https://scholar.google.com/)

Provides a broad search for scholarly literature across many disciplines and sources: articles, theses, books, abstracts from academic publishers, professional societies, online repositories, universities and other websites.

We also searched the bibliographies of retrieved systematic reviews.

## Inclusion criteria

We included prospective or retrospective observational studies, including case series and ecological designs, or interventional studies including randomised trials and clinical reports, outbreak reports, case-control studies and experimental studies. Studies incorporating models to describe observed data were included; however, studies reporting solely predictive modelling will be excluded. Single case reports were excluded, as no case report would have information on secondary cases. Studies not reporting data by symptom status were excluded as those with a single observation point. This is because a single observation or inadequate follow up cannot distinguish between presymptomatic, symptomatic and asymptomatic cases.

Studies were included if they reported the following information:

***Population:*** people exposed to SARS CoV-2 within 2-14 days (incubation time) of close contact or suspected community or institutional exposure to index asymptomatic (at the time of observation) infected individuals, as defined in the study.

***Reference:*** secondary case infected based on fulfilling a confirmed or probable case definition

***Target:*** level 3 / level 4 evidence with confirmed transmission outcome^10^.

We modified our original protocol to define the contribution to our evidence base from included studies. First, we included all studies satisfying our overall inclusion criteria. Second, for assessing the chain of transmission (question 4), we included those studies which identified index cases and estimated the potential for secondary transmission and thus allowed for analysis. The data extracted to document transmission included i) documentation of the likelihood of transmission; ii) presence of replicating virus and/or documentation of phylodynamics (genetic sequence lineage); and/or iii) adequate follow-up and reporting of symptoms and signs. [See box 1 for explanation]. For relevant studies, where necessary, one review author wrote to the corresponding author (with one reminder if necessary) to request further details.

### Box 1

#### i. Was transmission documented?

i1. Was the chain of transmission adequately described and reported?

Demonstrable and replicable chain of transmission (Gwaltney’s postulates^11^).

- Viral growth at the proposed anatomic site of origin;
- Viral contaminant reaches portal of entry of new host;
- The results of the study are replicated independently based on the methods detailed in the first study.

The last item should be considered an ideal aim, as many transmission studies are one-off and observational. Outlying studies and those that reach different conclusions should be assessed to ascertain reasons for diversity.

i2. Are the circumstances of transmission adequately assessed and reported?

- Context (exposure takes place).
- Environment (temperature, relative humidity, air exchanges, ultraviolet light etc.).
- Route if known - report if multiple possible routes are entertained or cannot be ruled out.
- Circumstances of exposure, sample collection and symptoms onset are reported.

#### ii. Were viable replicating viruses and/or phylodynamics documented?

ii1. The presence of a viable replicating virus with phylodynamics compatible with hypothesised source ascertained.

- Cq, Ct, Log concentration or number of copies are assessed and reported.
- Observed structural changes in host cells caused by the viral invasion that leads to visible cell lysis and/or other cytopathic phenomena or equivalent in culture.
- Evidence of virus replication consistent with expected growth kinetics in appropriate cell lines.
- Testing for evidence of contamination by other infectious agents.

ii2 Serial Culture adequately described and reported

- Techniques measuring viral infectivity using appropriate cell lines (e.g. viral plaque assay, TCID50 and immunofluorescence).
- For guidance, see section 11-b of *Use of cell culture in virology for developing countries in the South-East Asia Region. New Delhi: World Health Organization, Regional Office for South-East Asia; 2. Licence: CC BY-NC-SA 3.0 IGO*.

ii3 Genome sequencing adequately described and reported [based on WHO Genomic sequencing of SARS-CoV-2: items^12^]

- Genome sampling strategies and study design are considered and reported, including the risk of cross-contamination [item 6.1].
- The location of sequencing was appropriate [item 6.3.1].

iii. Was there a precise definition of symptoms and signs used, and was follow-up adequate?

- Could the patient flow or data collection methods introduce bias, and were measures to mitigate the bias introduced?
- Inadequate follow-up may misclassify pre-symptomatic individuals^3^. A follow up period is required to assess the presence or absence of symptoms and signs.
- An assessment of other underlying reasons for symptoms and signs should be applied in all cases

A reassessment of symptoms and signs should be recorded by another interviewer in a proportion of the cases as a data quality check.

## Quality Assessment

There are no formal quality assessment and reporting criteria for transmission studies, a situation reminiscent of the early days of Evidence-Based Medicine. Some authors have adapted observational checklists to assess quality^9^. However pre-existing tools and adaptations do not adequately account for the biases that might influence the understanding of the chain of transmission and the need to obtain microbiological as well as clinical confirmation of transmission. As this aspect is even more critical in the case of asymptomatic transmission, we created the list in Box 1, included only level 3 / level 4 evidence^10^ and further broke down the contribution of each study. Once we had assessed the items in Box 1, we looked at how each study defined, assessed and reported the absence or presence of signs and symptoms of Covid-19 in the respective populations.

We consider the assessment for the precise definition of symptoms and signs (set in the knowledge when each study was carried out) used and whether the follow up was adequate (item iii) as essential to determining the transmission of SARS-CoV-2 from pre and asymptomatic infected individuals. Incorporating the latter two criteria in the methods was considered essential to minimize bias by applying objective defined symptom criteria, a defined period of symptom assessment both before and after the testing period, and the method of ascertainment. To assess the quality of the symptom methods, one reviewer extracted the information on the symptom methods, the criteria to define those classified as symptomatic, asymptomatic and presymptomatic. We also included the author responses to requests for additional information in our assessment of bias. One clinical reviewer categorized the potential for bias as high, moderate, or low, which was independently checked by a second clinical reviewer. Reasons for the bias assessment for each study were also recorded.

## Data extraction

Search yields were screened in duplicate, and included study data were extracted into templates that included study characteristics, methodological aspects of studies and a summary of the main findings. Two reviewers also extracted data on the inclusion criteria for the review and the data for the transmission analysis. We followed PRISMA reporting guidelines as indicated for systematic or scoping reviews where applicable^13^. Data extraction was performed by one reviewer and independently checked by a second reviewer. In cases of disagreement, a third author arbitrated.

## Data synthesis and reporting

Outcomes of interest are listed in the inclusion criteria. We summarised data narratively and reported the outcomes as stated in the paper, including quantitative estimates and measures of dispersion where feasible and relevant. We reported subgroups of results by setting where appropriate and where sufficient details were provided in the included papers (e.g., care homes, detention centres, educational settings, hospitals, households, and passengers). We wrote to all study authors for clarification of methods and data. To assign the likelihood of transmission, two reviewers (CJH, TJ) used the existing <u>WHO Uppsala Monitoring Centre</u> (UMC) framework standardised case causality assessment and adapted it for SARS-CoV-2 transmission (Appendix C). The causality categories included certain, probable/likely, possible, unlikely or unclear. Clarification was sought from study authors, and where there was disagreement, consensus was reached by discussion.

## Results

The literature searches identified 444 records for screening for inclusion in this review (Figure 1): 388 studies were excluded after title and abstract screening. A further 39 were excluded on full-text analysis (see Figure 1 for the reasons for exclusion).

**Figure 1.**
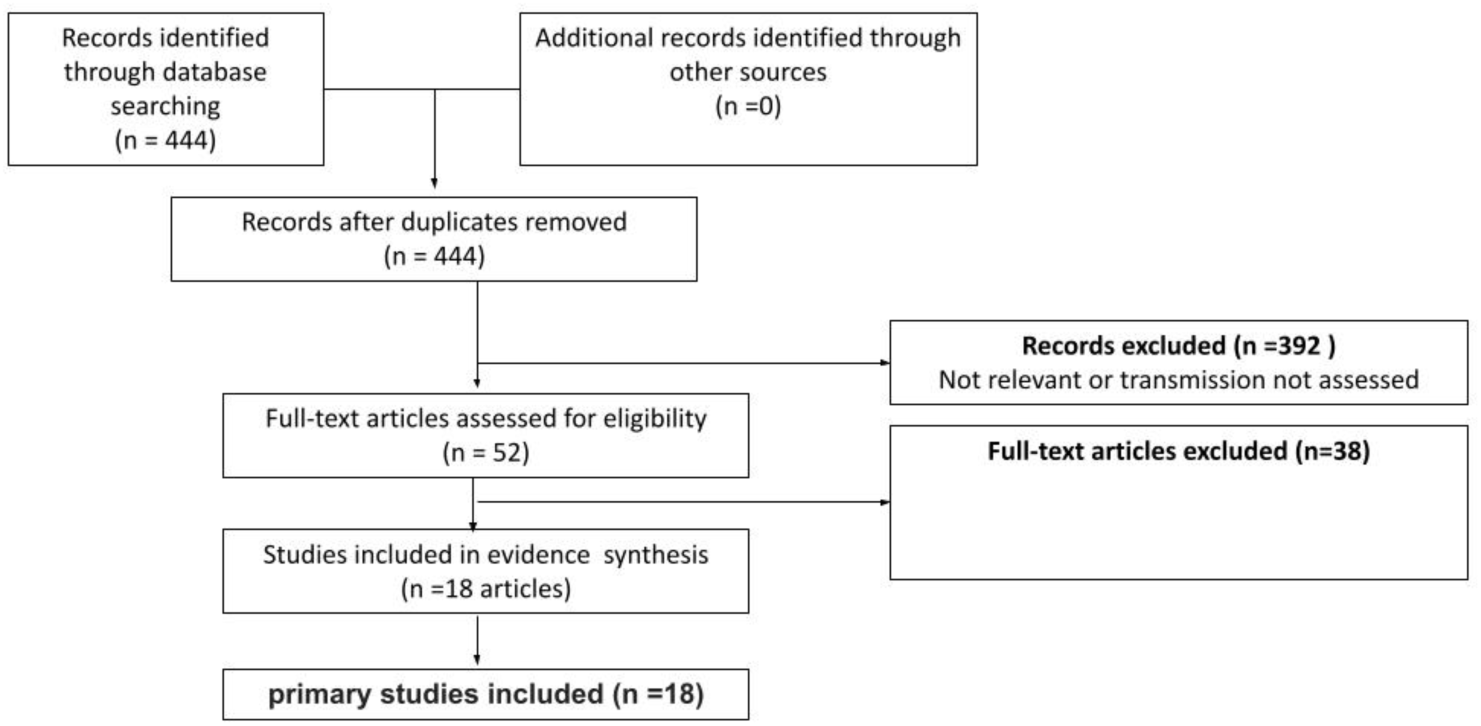
Flow Chart for Asymptomatic Transmission

The list of excluded studies is available on request from the corresponding author. Finally, we included 18 studies in the review (See Appendix A for included studies references list). We then wrote to 17 corresponding authors (of 18 papers) and received 17 responses. After receiving the responses, we included 17 studies in the review. See Appendix B for the characteristics of included studies.

Of the 17 included studies, seven were done in the USA (Arons, Gettings, Hershow, Lewis, Pray, Surie, Wallace), 6 in Europe (Borges, de Laval, van Hensenbergen, of which three were in London, UK-Cordery, Jeffery-Smith, Taylor), two in Canada (Ferreira, Soto), one in Japan (cruise ship) (Murata) and one in Australia (flight) (Speake). Most studies were completed in 2020, with 11 in the first half of the year (Arons, de Laval, Ferreira, Jeffery-Smith, Lewis, Murata, Soto, Speake, Taylor, van Hensenbergen, Wallace) and four in the second half (Borges, Cordery, Pray, Surie). Two studies were done between December and January 2021 (Gettings, Hershow).

Four studies were done in long term care facilities (Arons, Jeffery Smith, Surie, van Hensenbergen), and one among patients in a hospital (Borges). Three studies were based in schools with or without associated households (Cordery, Hershow and Gettings). Other settings included an emergency childcare centre (children and staff) (Soto), a detention centre (Wallace), passengers on a flight (Speake), passengers disembarking from a cruise ship (Murata), staff in an army barracks (Taylor), a military facility (de Laval), healthcare workers in a hospital (Ferreria), households (Lewis) and a university campus (Pray). Included studies had varied designs, including cross-sectional, repeat surveys, symptom-responsive screening designs, and studies addressing varying research questions.

Nasopharyngeal/oropharyngeal/throat samples were collected and tested by RT-PCR in all studies except Hershow, a schools based study that used saliva samples tested by RT-PCR (Hershow). Samples were subjected to viral culture in 9 studies (Arons, Cordery, Lewis, Murata, Pray, Speake, Surie, Taylor, Wallace), and genome sequencing was applied in 10 (Borges, Ferreira, Gettings, Hershow, Jeffery-Smith, Soto, Speake, Taylor, van Hensenbergen, Wallace). In addition, two studies performed serology to assess immunological response (de Laval, Ferreira).

## Quality assessment

Studies with a documented index case (or cases), with confirmation of the index case by serial viral culture, or evidence from Ct/Cq/increasing viral load, or evidence from a comprehensive epidemiological investigation on transmission coupled with genomic sequencing, were few (possibly only Cordery, Wallace, Lewis, Soto).

Appendix C shows the quality of symptom assessment for each included study. While each study has strong methodological aspects, the current lack of standardized methodology and clear reporting criteria promotes substantial methodological variation in transmission studies. Examples include differences in measurement thresholds (e.g., temperature above, 37.5, 37.8 or 38C), symptoms collected (the covid list of symptoms has expanded over time), the mode of collection (self-reported, use of a checklist or structured questionnaire, interview or chart review or a combination of methods). Fourteen studies reported the criteria for symptomatic status, four (Arons, Ferreira, Murata, Wallace) reported the criteria for asymptomatic assessment and [Arons, Ferreria, Lewis and Wallace] reported the criteria for presymptomatic status.

Before and after a positive test, the timings of the data collection are crucial for determining symptomatic status. In Surie 2021 (moderate bias), each participant was followed for 42 days from enrollment. For the first 21 days, participants visited every three days; for the next 21 days, participants visited weekly. Symptom assessment and medical chart review were repeated at each visit. At enrollment and visits, participants were interviewed by project staff about symptoms. Symptoms before enrollment were also assessed by healthcare personnel at the facility using a standard symptom list. At the first PCR-positive test, 11 (65%) participants did not report any COVID-19 symptoms, but all became symptomatic (one on day 25). In Speake, of 29 passengers who were subsequently identified as PCR-confirmed SARS-CoV-2 linked to the flight, seven had symptoms, but no criteria or follow-up was provided for the 22 passengers presumed asymptomatic. At the time (March 2020), people with no symptoms were not tested because of a lack of availability. This means that those appearing as asymptomatic on the flight could have been pre-symptomatic; incomplete follow-up does not allow a complete picture to emerge.

A checklist or symptom assessment form was used by eight studies (Arons, Hershow, Lewis, Pray, Taylor, Wallace, Cordery, de Laval); interviews by six (Arons Jeffery-Smith, Surie, van Hensbergen, Wallace, de Laval) and one study used text-message based symptom monitoring (Gettings). In Arons et al., a standardized symptom-assessment form was completed by nurses and symptoms present during the preceding 14 days were recorded based on interview and review of medical records. In Wallac,e the investigators administered a structured survey, detainees completed a self-administered, paper-based questionnaire of symptoms in the preceding two months and two weeks, and on the day of each subsequent test, detainees received an abbreviated self-administered, paper-based questionnaire of symptoms experienced since their last test. Responses were verbally verified, and medical history data were abstracted from medical records. Overall, fifteen studies were classified as high risk and three studies at moderate risk of symptom ascertainment bias. In practice, this assessment means that high-risk studies may be less likely to distinguish between pre-symptomatic and asymptomatic status.

## Proportion Asymptomatic and Presymptomatic at the time of testing

Figure 2 shows the fifteen studies that reported asymptomatic cases (n=304 participants). Table 1 shows that the proportion asymptomatic at the time of testing varied significantly across studies (range 12.5%% to 100%) depending on the setting, case ascertainment methods, and population.

**Figure 2.**
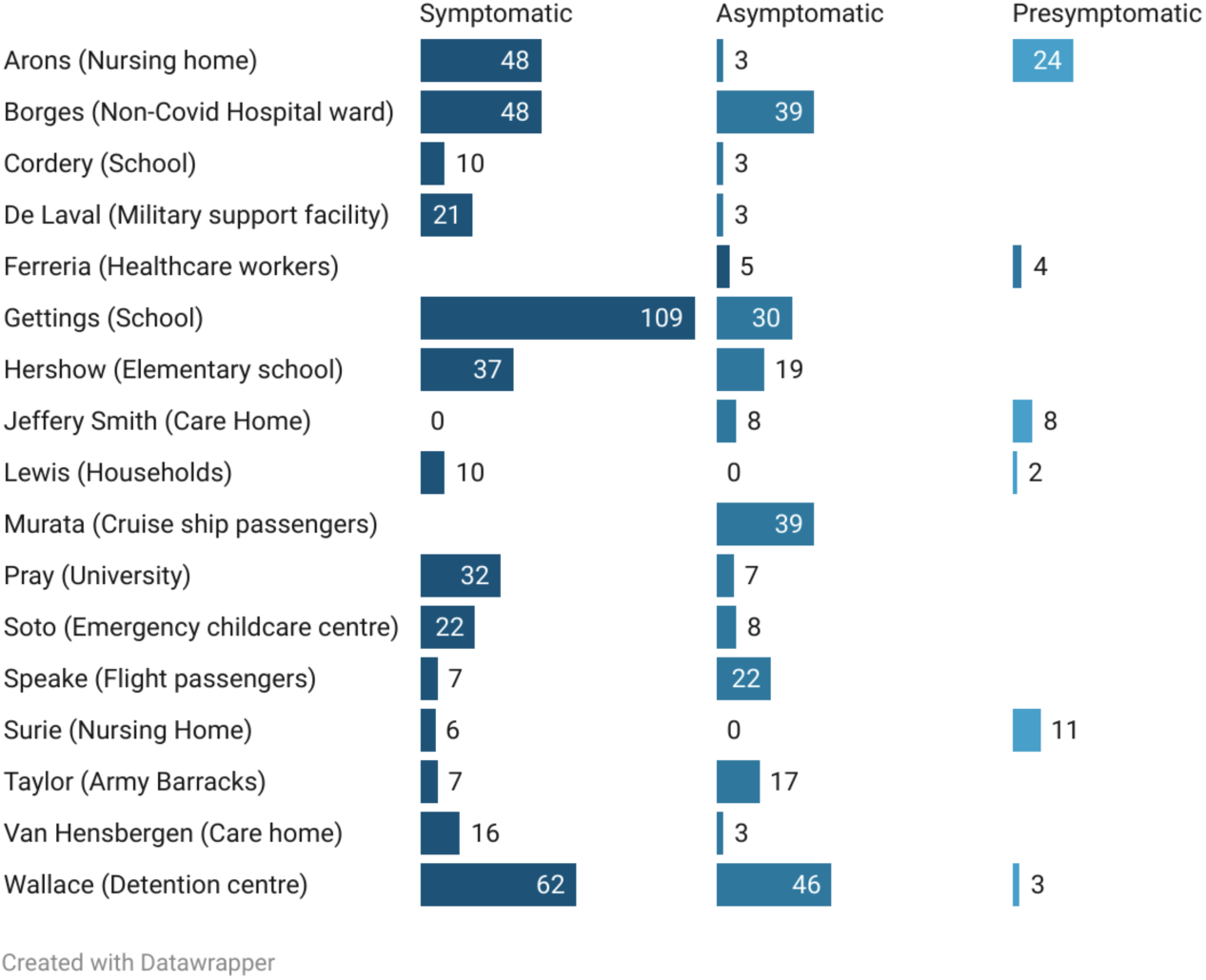
Number of asymptomatic, presymptomatic and symptomatic infected individuals from 17 studies.

**Table 1.**
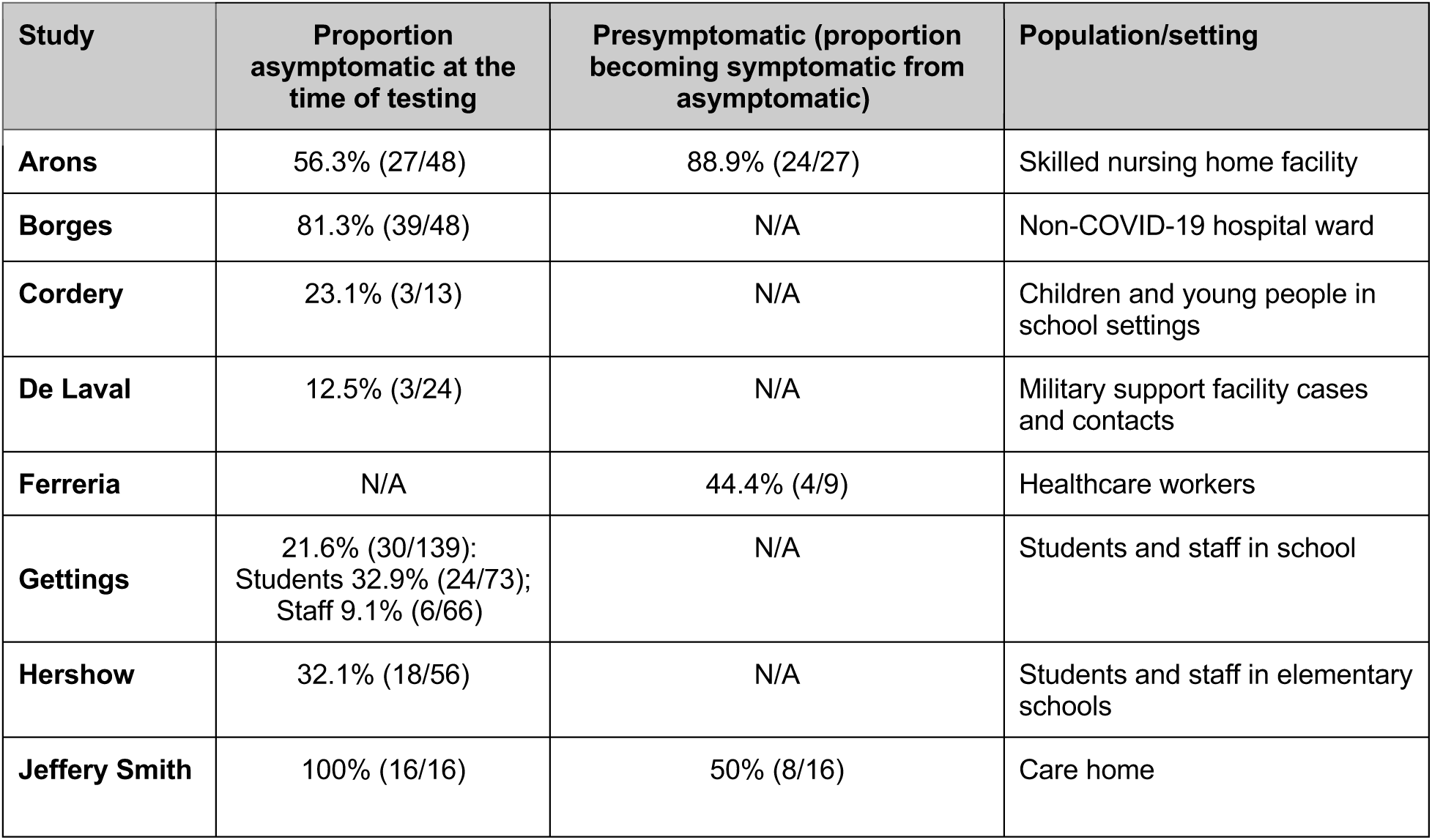
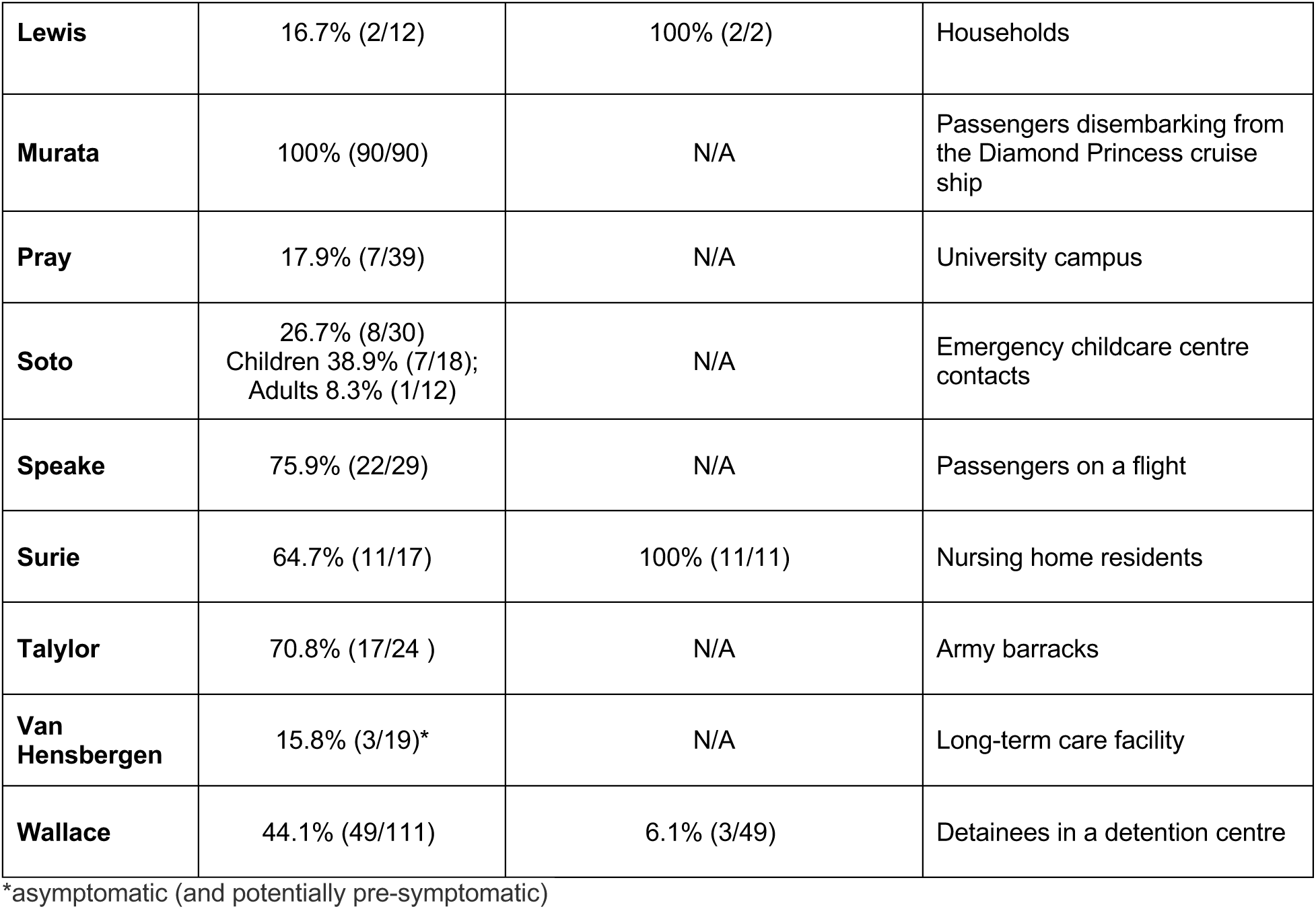
Numbers of Symptomatic, Asymptomatic and Presymptomatic cases at the time of initial testing.

In Gettings, the proportion asymptomatic was higher in students than staff: 32.9% vs 9.1% respectively, RR=3.6 (95% Confidence Interval, 1.58 to 8.30, p=0.002). In Soto, children were more likely to be asymptomatic: 38.9% vs 8.3%. Although this result was not significant, RR = 4.67 (95% CI, 0.65 to 33.30, p=0.12), this observation is consistent with a series from Manitoba, Canada, of 207 children aged up to 17, where infected children were significantly more likely than adults of being asymptomatic (37.8% [66/175] vs 7% [9/130]; p< 0.001, to have a significantly higher cycle threshold and to be significantly less likely to shed infectious virus^14^.

### Presymptomatic

Table 1 also shows the six studies that reported presymptomatic cases (n=50 participants) [Arons, Ferreria, Jeffery Smith, Lewis, Surie and Wallace]. Three of these were done in nursing/care home facilities and reported high rates of presymptomatic: 50 (8/16) in Jeffrey Smith, 89% (24/27) in Arons and 100% (11/11) in Surie Figure 2).

In Jeffrey Smith, the authors report atypical symptoms, which included but were not restricted to new confusion, reduced alertness, fatigue, lethargy, reduced mobility and diarrhoea. Arons reported viable SARS-CoV-2 was isolated from specimens of asymptomatic and presymptomatic residents but provided no level of quantification of the virus. Chronic symptom escalation in the elderly was considered a subjective assessment, and it was noted the elderly might present with more subtle symptoms requiring clinical assessment for detection. In Surie, patients often had difficulty distinguishing acute from chronic symptoms, especially for nonspecific symptoms such as fatigue and myalgia, which may account for the high percentage of recorded presymptomatic residents.

In Ferreira, the absence of symptoms was confirmed for all participants at the time of testing, and healthcare workers were followed for two weeks. In Lewis, two individuals were identified as testing positive before symptom onset. In Wallace, asymptomatic detainees reported no symptoms throughout observation (2 weeks). Presymptomatic cases reported >1 symptom after their first positive test with no previous symptoms.

## Infectious Status of asymptomatic and presymptomatic

Nine studies [Arons, Cordery, Lewis, Murata, Pray, Speake, Surie, Taylor and Wallace] assessed the infectious status of asymptomatics (n =7) and presymptomatics (n=4). Six asymptomatic studies and four presymptomatic studies demonstrated viral culture. Arons and Wallace assessed both asymptomatic and presymptomatics (see Table 2).

**Table 2.** Infectious status of asymptomatic and presymptomatic. Key: CPE=cytopathic effect.

| Study | Asymptomatic infectious | Pre symptomatic infectious | Culture method |
| --- | --- | --- | --- |
| Arons | 1/3 remained asymptomatic | 17/24 were presymptomatic | RT-PCR Positive specimens were inoculated in Vero-CCL-81 cells. Cells showing the cytopathic effect were used for SARS-CoV-2 RT-PCR to confirm isolation and viral growth in culture. |
| Cordery | No viral growth observed |  | Samples with high viral load (Ct value <30) were inoculated into Vero cells for the culture of infectious virus cell culture, as previously reported. |
| Lewis |  | No culturable and potentially infectious virus could be isolated | Viral cultures were performed at CDC.* |
| Murata | 7/39 asymptomatic carriers | | SARS-CoV-2 carriers who had ≥ two positive RT-PCR results at the hospital were analyzed using VeroE6/TMPRSS2 cells for the presence of viable viruses. Cells were checked every five days to see if they exhibited CPE. |
| Pray | Six asymptomatics out of 32 culture positives. |  | Virus culture was attempted on all antigen-positive or RT-PCR-positive specimens. Specimens were used to perform a limiting-dilution inoculation of Vero CCL-81 cells, and RT-PCR tested cultures showing evidence of CPE for SARS-CoV-2 RNA.*** |
| Speake | 1/6 asymptomatic passengers infectious in the mid-cabin and 3/5 in the aft cabin |  | Clinical specimens were inoculated in triplicate wells with Vero-E6 cells at 80% confluency, incubated at 37°C in 5% CO<sub>2</sub>, and inspected for cytopathic effect daily for up to 10 days. In-house PCRs confirmed the identity |
| Surie |  | One participant became symptomatic on day three after a positive RT-PCR test and was culture-positive on the initial test day. One severely immunocompromised participant shed replication-competent virus for 19 days from the positive test (17 days from symptom onset)**** | All oropharyngeal or anterior nares specimens with a positive RT-PCR were submitted for viral culture and inoculated in Vero CCL-81 cells and observed for CPE daily. When CPE was observed, the presence of SARS-CoV-2 was confirmed by RT-PCR. |
| Taylor | Live virus recovered in 6/7 (86%) at test visit 1.** |  | SARS-CoV-2 positive samples with a Ct <35 were subjected to virus isolation on Vero E6 cells swabs, and CPE confirmed virus detection up to 14 days after inoculation |
| Wallace | 12/52 asymptomatic culture positive | 2 of 3 pre-symptomatics were culture-positive | All positive samples were shipped to CDC for viral culture by using Vero-CCL-81 cells. The positive viral culture was confirmed in cells that showed a cytopathic effect by using rRT-PCR. |

\*For further details, see: Severe acute respiratory syndrome coronavirus two from a patient with coronavirus disease, United States. Emerg Infect Dis. 2020;26:1266–73.10.3201/eid2606.200516
\*\* One individual developed symptoms on the day of testing
\*\*\*Viral recovery was defined as any culture in which the first passage had an N1 Ct at least twofold lower than the corresponding clinical specimen.
\*\*\*\* data in Figure 4 in the paper identifies the immunocompromised patient as participant Q and the individual who became symptomatic on day three as participant K.

## Duration of infectiousness in asymptomatics and presymptomatics

### Asymptomatic

Murata examined SARS-CoV-2 cell infectivity in samples longitudinally obtained from asymptomatic carriers, and viable viruses from seven were isolated predominantly within seven days after the initial positive PCR test, except for one person who shed viable virus until day 15 (see appendix B). The specimen at day 15 (Ct 30.3) was from a 70-year-old Japanese female with a history of diabetes mellitus and hypertension who had prolonged RT-PCR positivity > 21 days.

### Presymptomatic

In Arons, 27 residents were classified as asymptomatic (15 reported no symptoms and 12 stable chronic symptoms). In the seven days after their initial positive test, 24 asymptomatic residents (89%) had onset of symptoms. The median time to symptom onset was four days (interquartile range, 3 to 5).

Lewis did not observe infectiousness in the two presymptomatic individuals. The 33-year-old woman (case 02-01) reported symptoms the day after a positive test, and in a 7-year-old girl, symptoms were reported after two days (case 02-03). In Surie, 9/17 participants (53%) had replication-competent virus isolated. One severely immunocompromised participant shed replication-competent virus for 19 days from the positive test (17 days from symptom onset) One became symptomatic on day three after a positive RT-PCR, test having been culture positive on the initial test day. The patient was hospitalized on day five and died.

## Relationship between infectiousness and PCR cycle threshold in asymptomatics and in presymptomatic

### Asymptomatics

The median CT of culture-positive individuals in Murata et al. were significantly associated with isolation of viable virus Ct 24.6 (IQR, 20.4-25.2) vs culture-negative Ct 35.9 (IQR, 33.5-37.1), P< 0.001. In Wallace, Ct for symptomatic (median 32.7, range 19.7-36.3) were comparable with asymptomatic (32.9, 19.8-36.9). The Median CT of culture-positive was 24.4 (IQR, 21.5-28.0; range, 19.8-33.7), and in the two culture-positive pre-symptomatics, Cts were 20 and 31.1). In Cordery, viral loads were reported as low in 2/3 of the cases (E gene Ct 34.5 and 35.6). In one case, the initial Ct was 26.3 that fell to 22.3 on day 4 (suggesting infectiousness) before increasing to 28.2 by day 8.

### Presymptomatic

In Lewis, the two presymptomatic household members’ initial ‘viral shedding’ corresponded with medium or high Ct values (1–2 days before symptom onset). Symptom onset in one patient (case 02-01) was associated with progression from a high Ct (> 30) to a medium value (Ct 20-30); symptom onset led to progression to a low value, < 20 suggesting active viral replication. In the other case (case 02-02, girl aged 7), the Ct remained in the range 20-30. Both cases reported high Cts (>30) on day 14. In Surie, replication-competent viruses could not be cultured above a Ct of 29. In Arons, the Ct values by symptom status were similar (asymptomatic, 25.5; presymptomatic, 23.1; atypical symptoms, 24.2; typical symptoms, 24.8).

## What is the evidence of a chain of transmission that establishes asymptomatic and presymptomatic transmission of SARs-CoV-2?

We used the existing WHO UMC framework and adapted it for SARS-CoV-2 transmission to assign the likelihood of transmission. There is an inevitable element of subjectivity which is why we report the rationale for the categorisation.

### Asymptomatics

Nine studies provided insufficient information [Borges, Ferreira, Francis, Gettings, Murata, Pray, Soto, Speake and Van Hensenbergen] to contribute to assessing the chain of transmission and were therefore classified as unclear. The classification is based on information from the paper and correspondence with authors. *Unclear* was assigned when a reasonable hypothesis was not formulated or tested or when reliance on consensus genome sequencing was the main component of transmission, but there was no ability or attempt to exclude other sources of infection.

Table 3 includes the six studies (Cordery, De Laval, Hershow, Jeffery Smith, Taylor and Wallace) that classified the probability the study contributes to the chain of asymptomatic transmission of SARs-CoV-2. Three studies were judged as possible (De Laval, Hershow, Jeffery Smith, and three as probable/likely - Cordery, Taylor and Wallace). In Wallace, 46 individuals were reported as asymptomatic, but the methodological limitations mean they could have also been presymptomatic.

**Table 3.** Evidence of the chain of transmission that establishes asymptomatic transmission of SARs-CoV-2.

| **Study** | **Probability** | **Reason** |
| --- | --- | --- |
| Cordery | **Probable/Likely** | Three asymptomatic cases were detected in week 2 of screening. There is no evidence of wider transmission among children remaining in school, except the one unexpected cluster of three asymptomatic cases in one school in the same class. In one of the asymptomatic cases, the viral load rose on repeat testing to >4 million copies per swab, and another asymptomatic household member was identified as infected (case A Cts: 26.3, 22.3, 28.2). However, the case remained asymptomatic despite viral shedding continuing for at least a week after an initial positive test. |
| De Laval | **Possible** | Three cases were asymptomatic. Contact tracing results did not identify any transmission from asymptomatic to symptomatic cases in this cluster. |
| Hershow | **Possible** | Low transmission in schools despite substantial community transmission. Among the five persons with school-associated cases, three persons were asymptomatic, and three were exposed to asymptomatic index patients; four cases were attributed to student-to-student transmission, and one was to student-to-teacher transmission. |
| Jeffery Smith | **Possible** | The finding of asymptomatic SARS-CoV-2 infection in care homes that did not report a single case of COVID-19 and genomic evidence of a |
|  |  | small cluster of staff and residents infected with the same SARS-CoV-2 lineage in care home F. It was not possible to extract direct information on transmission from identified asymptomatic index cases to contacts. |
| Taylor | **Probable/Likely** | There were 4 cases that all remained asymptomatic throughout with 0 base difference (genetically indistinguishable); these 4 cases had a link through one common workplace location. There were another 4 (different) cases, although two later developed symptoms also with 0 base difference; other than visiting the same shop and using a common entrance to the barracks, no common links in the workplace/barracks setting could be found. |
| Wallace | **Probable/Likely** | 12/52 asymptomatic had positive viral culture results. A large number of asymptomatic infections and shedding of replication-competent virus in asymptomatic participants. The phylogeny indicates within-dormitory transmission. Individuals are described as asymptomatic, but they could have also been presymptomatic. |

### Presymptomatics

Table 4 classifies the probability the study contributes to the chain of transmission that establishes presymptomatic transmission of SARs-CoV-2. Of the six studies, Ferreria provided insufficient information to assess the chain of transmission and was judged unclear. Of the five studies, two were assigned as possible [Jeffery Smith, Surie], one unlikely (Lewis) and two as probable/likely (Arons, Wallace). Two studies were classified as unlikely because another explanation (the likely symptomatic index) was more plausible.

**Table 4.** Evidence of a chain of transmission that establishes presymptomatic transmission of SARs-CoV-2.

| **Study** | **Probability** | **reasons** |
| --- | --- | --- |
| Arons | **Probable/Likely** | Viral growth was observed for specimens obtained from 17/24 presymptomatic residents; 24 presymptomatic residents had a median rRT-PCR Ct value of 23.1. More than half of residents with positive test results were asymptomatic at the time of testing and most likely contributed to the transmission. Staff and residents were being actively screened for signs and symptoms and either promptly isolated (residents) or excluded from work (staff) if any were present. |
| Jeffery Smith | **Possible** | The finding of asymptomatic SARS-CoV-2 infection in care homes that did not report a single case of COVID-19 and genomic evidence of a small cluster of staff and residents infected with the same SARS-CoV-2 lineage occurred in care home F. It was not possible to extract direct information on transmission from identified pre-symptomatic index cases to contacts. |
| Lewis | **Unlikely** | Five households enrolled. Eligibility entailed an identified positive index case resulting from testing due to symptom onset within each household; secondary transmission was observed in two households. WGS for the second household (HH 05-00 symptomatic) indicated the likely chain of transmission was from 05-00 and/or 05-03 (symptomatic) who had genetically identical infections and were exposed to the same community contact. |
|  |  | WGS indicates that the infections across all four household members in HH-2 were essentially genetically identical, suggesting that the index case, 02-00 (symptomatic), was transmitted to all remaining household members. |
| Surie | **Possible** | Whole-genome sequencing on eligible specimens (Ct <30) showed there were only 2-3 single nucleotide variant differences among the entire set of sequenced genomes, which implied they were likely from the same source and a single introduction the nursing home. It is not clear who is the source, and there were six symptomatic individuals. Thus, 9/17 participants (53%) had replication-competent virus isolated. The authors consider that the findings underscore the potential role of pre-symptomatic carriers in transmission. |
| Wallace | **Probable/Likely** | 12/52 asymptomatic had positive viral culture results. A large number of asymptomatic infections and shedding of replication-competent virus in asymptomatic participants. The phylogeny indicates within-dormitory transmission. Individuals are described as asymptomatic, but they could have been presymptomatic. |

## Discussion

Our review was designed to address the question of whether and how much transmission from individuals, either asymptomatic at the time of testing or remained so throughout the length of their positivity, was documented with epidemiological and laboratory evidence of high reliability. This was based on genome sequencing and/or viral culture of samples to indicate infectious potential and epidemiological tracing to identify onward transmission. Sequencing ascertained phylodynamics and lack of contamination or co-infection, while culture indicated whether the isolate could replicate and spread to infect other cells to perpetuate the process. In parallel, PCR positivity identified those who had been in contact with some of the antìgens. No single measure sufficed but taken together with clinical history, these four variables can narrow the transmission uncertainty. Despite many studies on transmission, we identified only 18 studies fitting our inclusion criteria. No one study provides the answer but taken together, the included studies stand witness to the efforts of their authors to address the issue of asymptomatic and presymptomatic transmission. A 100% response rate to reviewers’ queries is, to our knowledge, very unusual in systematic reviews^15^. The willingness shown by the corresponding authors of all our included studies in responding to all our queries and providing extra details and data should be capitalised on as the beginning of an international effort to standardise methods and reporting of viral transmission studies drawing together the epidemiological, clinical and virological strands.

Transmission studies are technically challenging to perform, especially amid a pandemic. It is also sometimes difficult to reconstruct events and to separate data for people with no symptoms at the time of the survey and the follow up of their contacts. In some cases, highly cited articles reporting asymptomatic spread turned later to be from an index case who was symptomatic at the time of exposure of contacts but who suppressed symptoms to carry on with her activities^16, 17^. For the Böhmer case, whilst additional information has become available post-publication, clarifying that the suspected index case did indeed suffer symptoms compatible with Covid-19^16^ there has not yet been a retraction or correction of that original letter which described, in its title, the index case as asymptomatic. This has allowed the misunderstanding of this case study to perpetuate a false ascertainment of asymptomatic transmission.

The answer to the question of what fraction of the total positives were asymptomatic throughout or only at the point of testing, could not be summarised in a single pooled estimate, given the wide differences in estimates that we have documented (12.5%% to 100% for asymptomatic at the time of testing and 6% to 100% of these being pre-symptomatic cases). Caution should, therefore, be applied to published pooled summary estimates, given the heterogeneity due to the setting, the methods of case ascertainment (including follow-up), the testing and the source population some of whom may have significant cognitive impairment or other factors precluding accurate and reliable symptom ascertainment.

The differences between asymptomatics and pre-symptomatics are also due to the methodologically weak practice of single point-in-time testing (with no or selective follow up) and use of a different sign or symptom definitions. In addition, some studies did not report what list (if any) was used. Several authors also reported difficulty in accurately assessing the symptom profile of elderly patients, suggesting that a more thorough clinical observation, careful observation for signs, and follow up are needed for accurate classification. The likelihood of symptom ascertainment bias is a quality criterion which we inserted after correspondence with the authors of all the studies included in this review and should be incorporated in any future work to identify the role of pre and asymptomatic subjects in the spread of SARS-CoV-2. As such, though, it is a post protocol item due to the evolution of our understanding of the subject matter and is prone to the problems associated with subjective assessments.

Single or point binary PCR testing alone (especially with no Ct reported) cannot give information on infectivity, as the work of Murata et al. shows. A follow up of up to 21 days after the first PCR test of 90 apparently asymptomatic cases from the Diamond Princess with repeated PCR tests, taken in conjunction with the clinical picture and reporting of serial (i.e. on the same subject) Cts, identified 39 true asymptomatic subjects with more than two consecutive or nonconsecutive positive PCR test results at the hospital - seven being potentially infectious.

The serial trend of Ct values, which is linked to the probability of culturing live viruses^14, 18^, is thus predictive of likely individual infectiousness, allowing adequate measures to be taken to interrupt the potential spread. We do not have sufficient data to explore the likelihood of infectiousness by age and risk group. Still, the evidence presented in this review shows that a large but variable percentage of asymptomatic subjects goes on to develop symptoms, which a single observation or test will not identify. Therefore, the labelling of a subject as “asymptomatic” based on a single observation is wrong and misleading.

We found evidence of likely infectiousness of both asymptomatics and pre-symptomatics, but we cannot quantify a percentage of infectiousness or contact risk for either category. In part, this is due to the variability of our included populations and perhaps also to varied testing and culture methods that cannot easily translate across different laboratories and cell lines. We exercised caution in comparing results from included studies as there are issues of comparability of cycle threshold values across different PCR ^19^. This again points to the requirement for a common approach.

We cannot be certain of the duration of infectiousness but note that there do not seem to be large differences in median cycle thresholds between potentially infectious asymptomatic and pre-symptomatic subjects; again, this is consistent with the observation from the Manitoba series ^14^.

Our assessment of the likelihood of transmission is based on what is essentially a headcount, and we phrased it in terms of likelihood, as an absolute proof is not easy to come by, especially in the absence of challenge studies that are difficult to do because of safety and ethics concerns ^20, 21^. The assessment was primarily based on our correspondence with the authors of the included studies and the additional information provided. There are limitations to this approach; however, our methods are designed to foster dialogue on the ideal methods for transmission and to provide a basis for a joint methods standard for assessing transmission and its reporting

A better understanding of transmission dynamics is essential for pandemic planning as with influenza and other respiratory agents. For example, if a substantial proportion of transmission occurs from individuals who at the time have no symptoms, some control measures such as quarantine and contact tracing might have doubtful value, especially if the duration of infectiousness is brief, but infectivity high, a pattern that fits with the infection of the youngest groups of infected people^22^.

The prolonged period of potential infectiousness reported by Murata et al.^23^ in one of the elderly asymptomatic subjects from the Diamond Princess cruise ship should also be investigated in other settings as it may explain the sudden onset of epidemic foci in nursing homes several days after admission of the last patient to the institution. Again, a single “entry” point test may not be enough. Furthermore, the potential for the elderly to present with more subtle symptoms that may go unrecognized and with a prolonged infectious period is important to understanding and containing outbreaks in care homes.

## Policy recommendations

This review includes a limited body of evidence on which to base public health recommendations. Therefore, research should be embedded into all public health interventions where substantial uncertainty exists, including the question of whether asymptomatic and presymptomatic individuals are important drivers of the transmission of SARS-CoV-2.

## Research recommendations

The observational nature of the studies’ design and the lack of universal methods and reporting standard hinder interpretation and point to the absolute need for such standards to be drawn and agreed upon by those interested in researching in vivo transmission of viral respiratory pathogens SARS-CoV-2 in particular. We recommend carrying out longitudinal follow-up studies of at least three weeks’ duration during epidemics with consolidated symptoms assessment testing and repeated serial PCRs to elucidate further and clarify the role of those who initially show no symptoms but are positive. At least one study per type of population or setting should be carried out. Graphic presentation of the results could be standardised. We found the reporting of the evolution of transmission in the study by Lewis et al. ^24^. particularly helpful and clear.

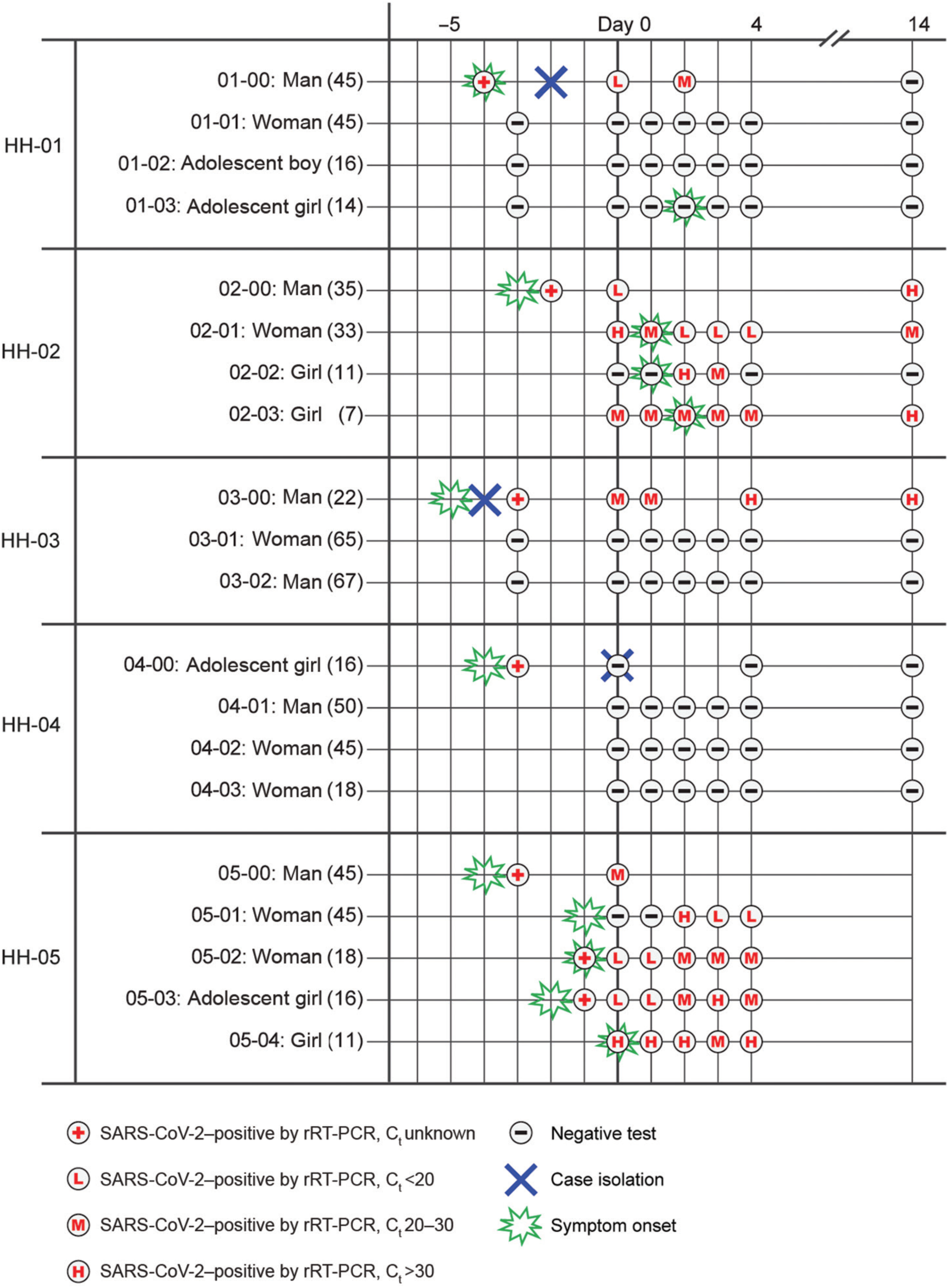

This is a reproduction of Figure 1 from the study by Lewis et al.^25^. The figure clearly illustrates the results of rRT-PCR for SARS-CoV-2, viral burden and symptom onset among index case-patients and SARS-CoV-2–positive and negative household contacts. Timelines of symptom onset and testing dates before and during the 15-day study period are reported by the household. Sex and age are reported on the left in brackets. Symptom onset date is only included for household members who tested positive at any time during the study period or for whom onset of symptoms consistent with coronavirus disease prompted an interim visit from investigators. Key: Ct = cycle threshold; HH = household; rRT-PCR = real-time reverse transcription PCR; SARS-CoV-2 = severe acute respiratory syndrome coronavirus 2.

## Conclusion

In summary, the results of the studies included here with more reliable testing and design, provide probable evidence of transmission of SARS-CoV-2 from presymptomatic and asymptomatic individuals. Single point in time estimates and binary PCR testing alone cannot provide reliable information on symptom status and information on infectivity. The number of studies and asymptomatic and presymptomatic cases eligible for inclusion was low, with more data and international standardisation of methods needed to reduce uncertainty further.

## Data Availability

Extraction sheets and list of excluded studies are available on request from the corresponding author

## Acknowledgements

This review would not have been possible without the input of Drs Vitor Borges, Victoria Chu, Victor Ferreira, Frank De Laval, John Jernigan, Jenna R. Gettings, Mitch Van Hensenberg, Nathaniel Lewis, Shamez Ladhani, Matthew Loose, Suzanne McEvoy, Ian Pray, Hannah Taylor, Aki Sakurai, Shiranee Shriskandan, Julio Soto, Diya Surie and Megan Wallace. We are grateful for their help.

## Conflict of interest statements

TJ was in receipt of a Cochrane Methods Innovations Fund grant to develop guidance on the use of regulatory data in Cochrane reviews (2015-018). In 2014–2016, he was a member of three advisory boards for Boehringer Ingelheim. TJ was a member of an independent data monitoring committee for a Sanofi Pasteur clinical trial on an influenza vaccine. TJ is occasionally interviewed by market research companies about phase I or II pharmaceutical products for which he receives fees (current). TJ was a member of three advisory boards for Boehringer Ingelheim (2014-16). TJ was a member of an independent data monitoring committee for a Sanofi Pasteur clinical trial on an influenza vaccine (2015-2017). TJ is a relator in a False Claims Act lawsuit on behalf of the United States that involves sales of Tamiflu for pandemic stockpiling. If resolved in the United States favor, he would be entitled to a percentage of the recovery. TJ is coholder of a Laura and John Arnold Foundation grant for development of a RIAT support centre (2017-2020) and Jean Monnet Network Grant, 2017-2020 for The Jean Monnet Health Law and Policy Network. TJ is an unpaid collaborator to the project Beyond Transparency in Pharmaceutical Research and Regulation led by Dalhousie University and funded by the Canadian Institutes of Health Research (2018-2022). TJ consulted for Illumina LLC on next generation gene sequencing (2019-2020). TJ was the consultant scientific coordinator for the HTA Medical Technology programme of the Agenzia per i Servizi Sanitari Nazionali (AGENAS) of the Italian MoH (2007-2019). TJ is Director Medical Affairs for BC Solutions, a market access company for medical devices in Europe. TJ was funded by NIHR UK and the World Health Organization (WHO) to update Cochrane review A122, Physical Interventions to interrupt the spread of respiratory viruses. TJ is funded by Oxford University to carry out a living review on the transmission epidemiology of COVID-19. Since 2020, TJ receives fees for articles published by The Spectator and other media outlets. TJ is part of a review group carrying out Living rapid literature review on the modes of transmission of SARS-CoV-2 (WHO Registration 2020/1077093-0). He is a member of the WHO COVID-19 Infection Prevention and Control Research Working Group for which he receives no funds. TJ is funded to co-author rapid reviews on the impact of Covid restrictions by the Collateral Global Organisation. TJ’s competing interests are also online https://restoringtrials.org/competing-interests-tom-jefferson

CJH holds grant funding from the NIHR, the NIHR School of Primary Care Research, the NIHR BRC Oxford and the World Health Organization for a series of Living rapid review on the modes of transmission of SARs-CoV-2 reference WHO registration No2020/1077093. He has received financial remuneration from an asbestos case and given legal advice on mesh and hormone pregnancy tests cases. He has received expenses and fees for his media work including occasional payments from BBC Radio 4 Inside Health and The Spectator. He receives expenses for teaching EBM and is also paid for his GP work in NHS out of hours (contract Oxford Health NHS Foundation Trust). He has also received income from the publication of a series of toolkit books and for appraising treatment recommendations in non-NHS settings. He is Director of CEBM and is an NIHR Senior Investigator.

DE holds grant funding from the Canadian Institutes for Health Research and Li Ka Shing Institute of Virology relating to the development of Covid-19 vaccines as well as the Canadian Natural Science and Engineering Research Council concerning Covid-19 aerosol transmission. He is a recipient of World Health Organization and Province of Alberta funding which supports the provision of BSL3-based SARS-CoV-2 culture services to regional investigators. He also holds public and private sector contract funding relating to the development of poxvirus-based Covid-19 vaccines, SARS-CoV-2-inactivation technologies, and serum neutralization testing.

JMC holds grants from the Canadian Institutes for Health Research on acute and primary care preparedness for COVID-19 in Alberta, Canada and was the primary local Investigator for a Staphylococcus aureus vaccine study funded by Pfizer for which all funding was provided only to the University of Calgary. He also received support from the Centers for Disease Control and Prevention (CDC) to attend an Infection Control Think Tank Meeting. He is a member of the WHO Infection Prevention and Control Research and Development Expert Group for COVID-19 and the WHO Health Emergencies Programme (WHE) Ad-hoc COVID-19 IPC Guidance Development Group, both of which provide multidisciplinary advice to the WHO and for which no funding is received.

JB is a major shareholder in the Trip Database search engine (www.tripdatabase.com) as well as being an employee. In relation to this work Trip has worked with a large number of organisations over the years, none have any links with this work. The main current projects are with AXA and Collateral Global. He worked on Living rapid literature review on the modes of transmission of SARS-CoV-2 (WHO Registration 2020/1077093-0) and is part of the review group carrying out a scoping review of systematic reviews and meta-analyses of interventions designed to improve vaccination uptake (WHO Registration 2021/1138353-0).

ECR was a member of the European Federation of Neurological Societies(EFNS) / European Academy of Neurology (EAN) Scientist Panel – Subcommittee of Infectious Diseases (2013-2017). Since 2021, she is a member of the International Parkinson and Movement Disorder Society (MDS) Multiple System Atrophy Study Group, the Mild Cognitive Impairment in Parkinson Disease Study Group, and the Infection Related Movement Disorders Study Group. She was an External Expert and sometimes Rapporteur for COST proposals (2013, 2016, 2017, 2018, 2019) for Neurology projects. She is a Scientific Officer for the Romanian National Council for Scientific Research.

IJO, EAS, and AP have no interests to disclose.

## Appendix A References of included studies (n=18)

## Appendix B Characteristics of included studies.

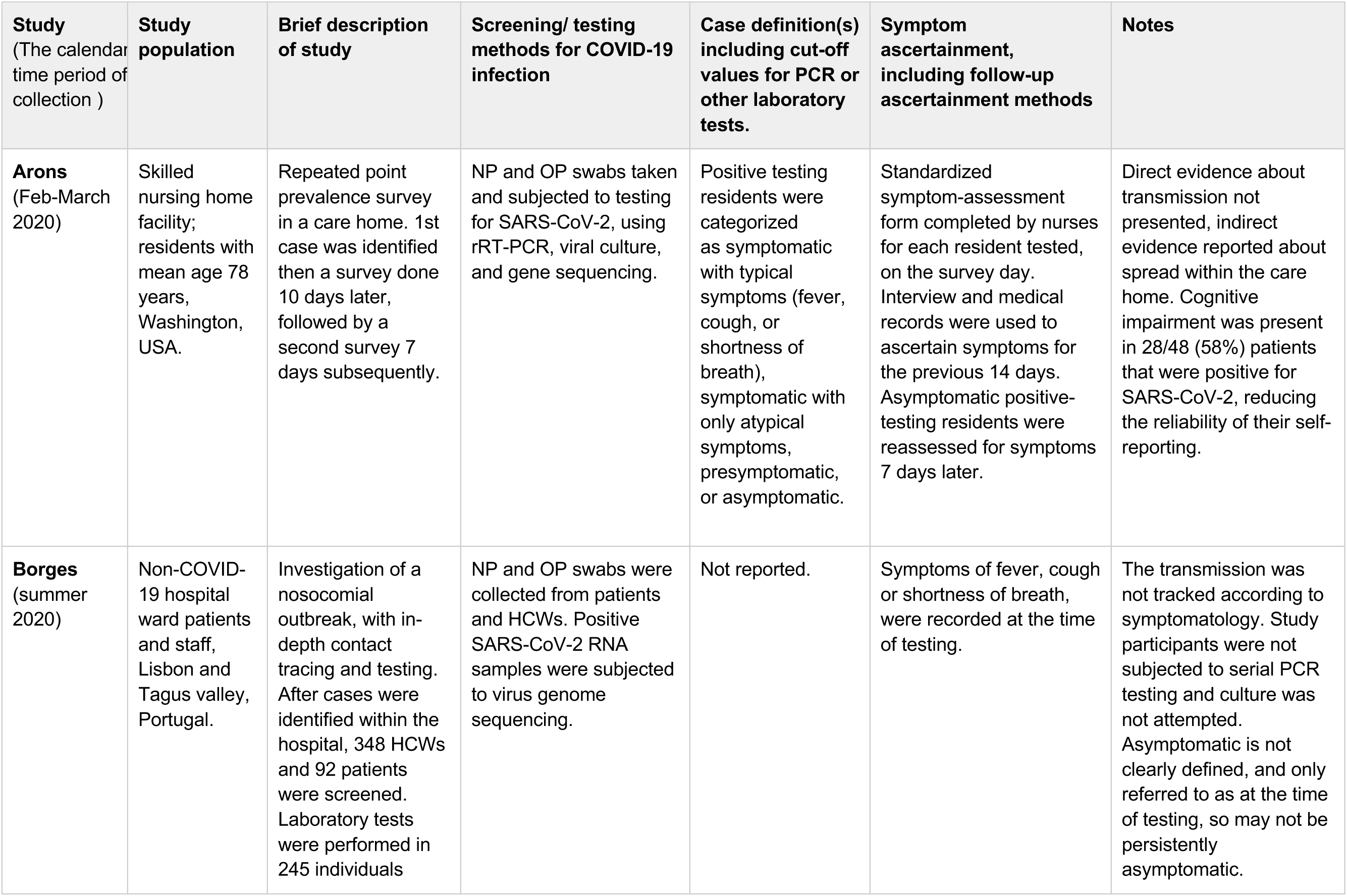

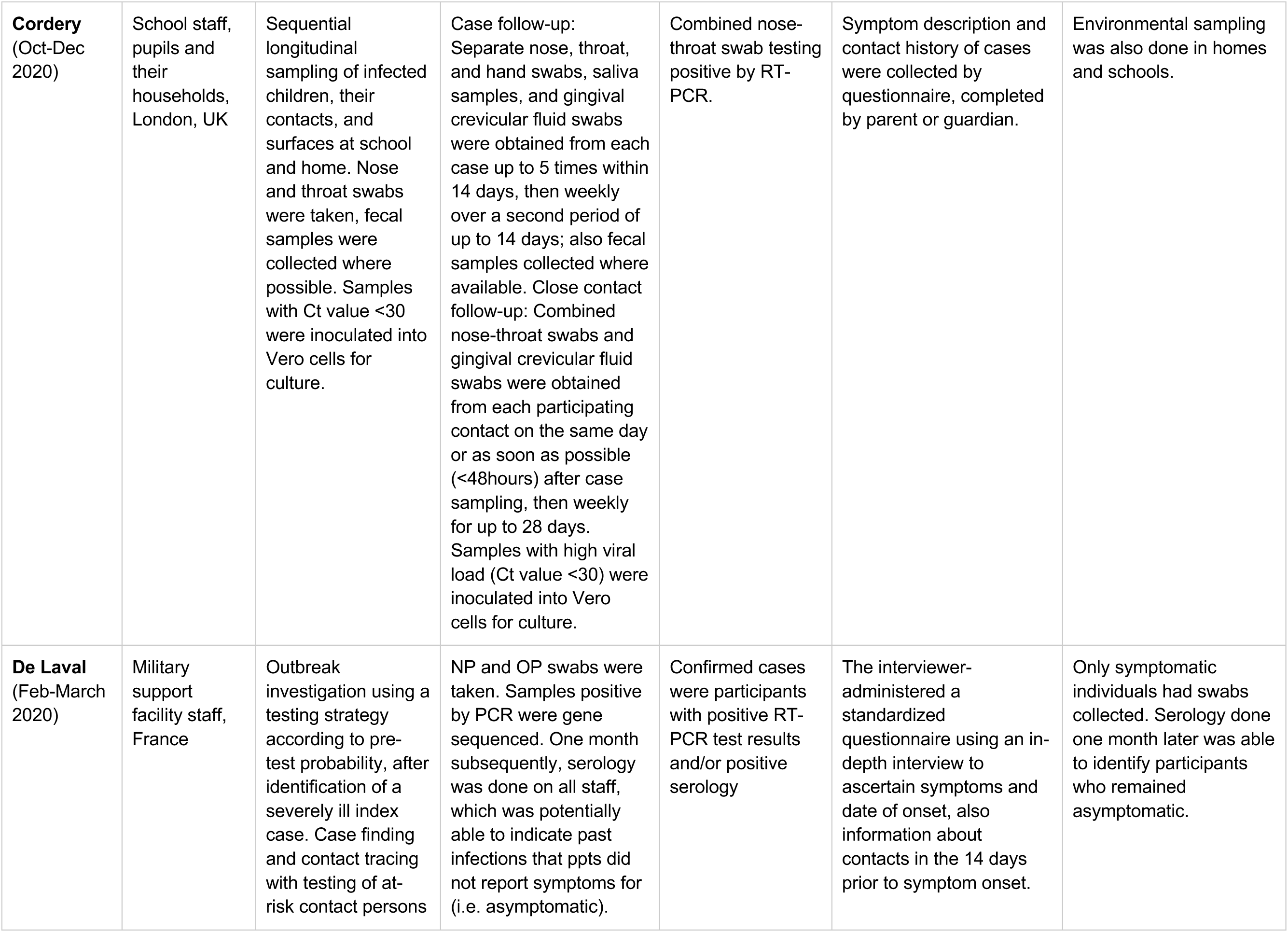

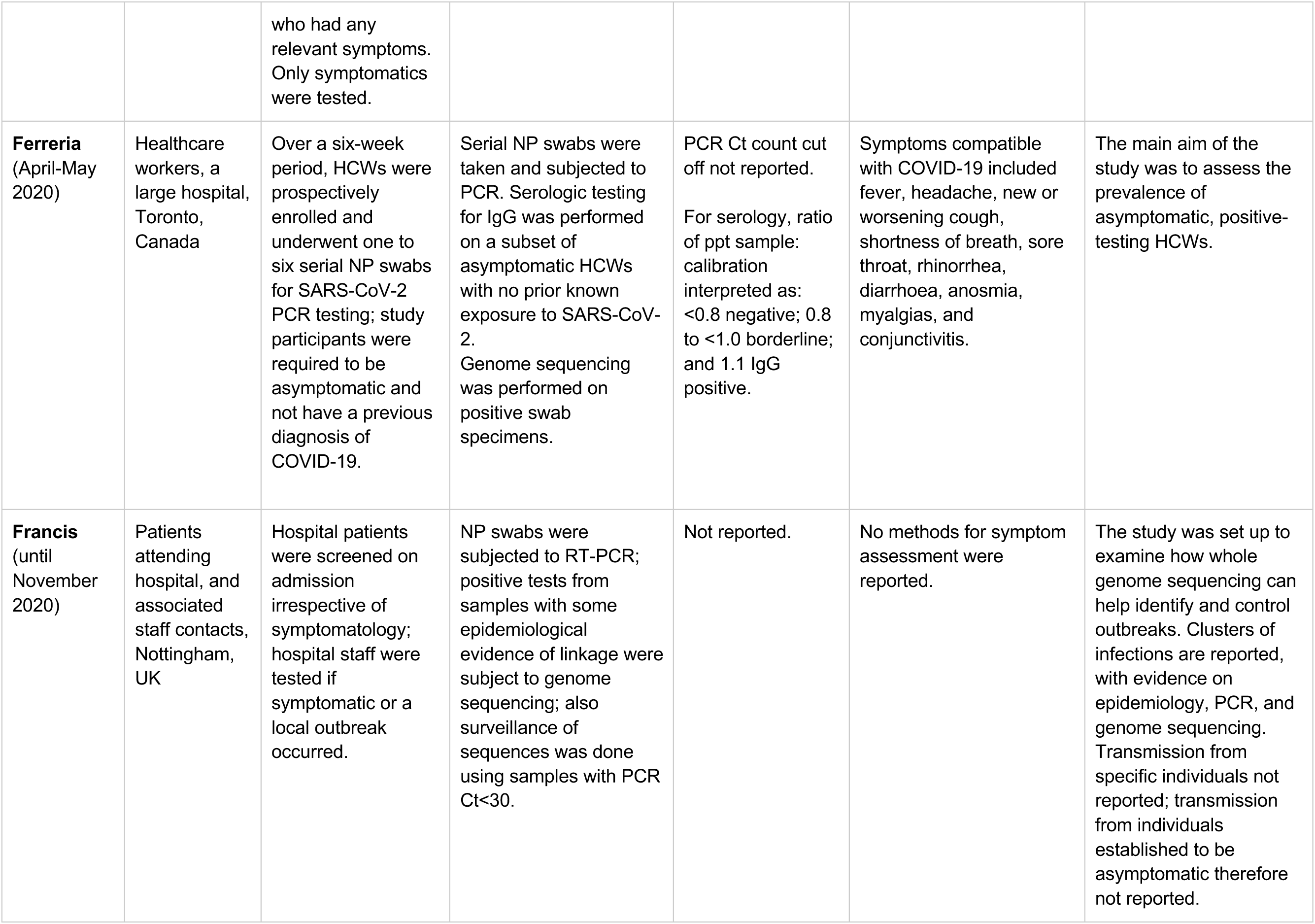

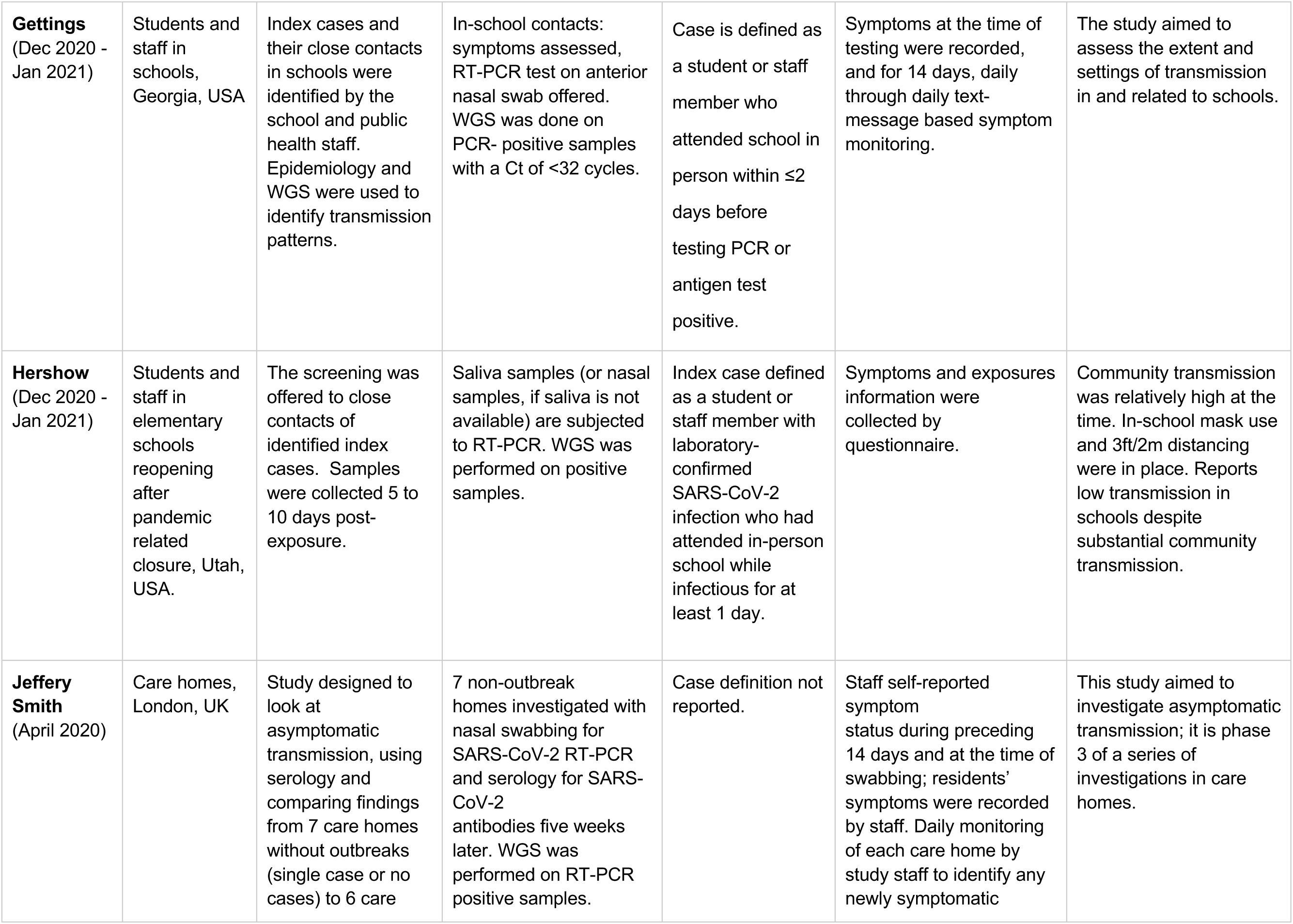

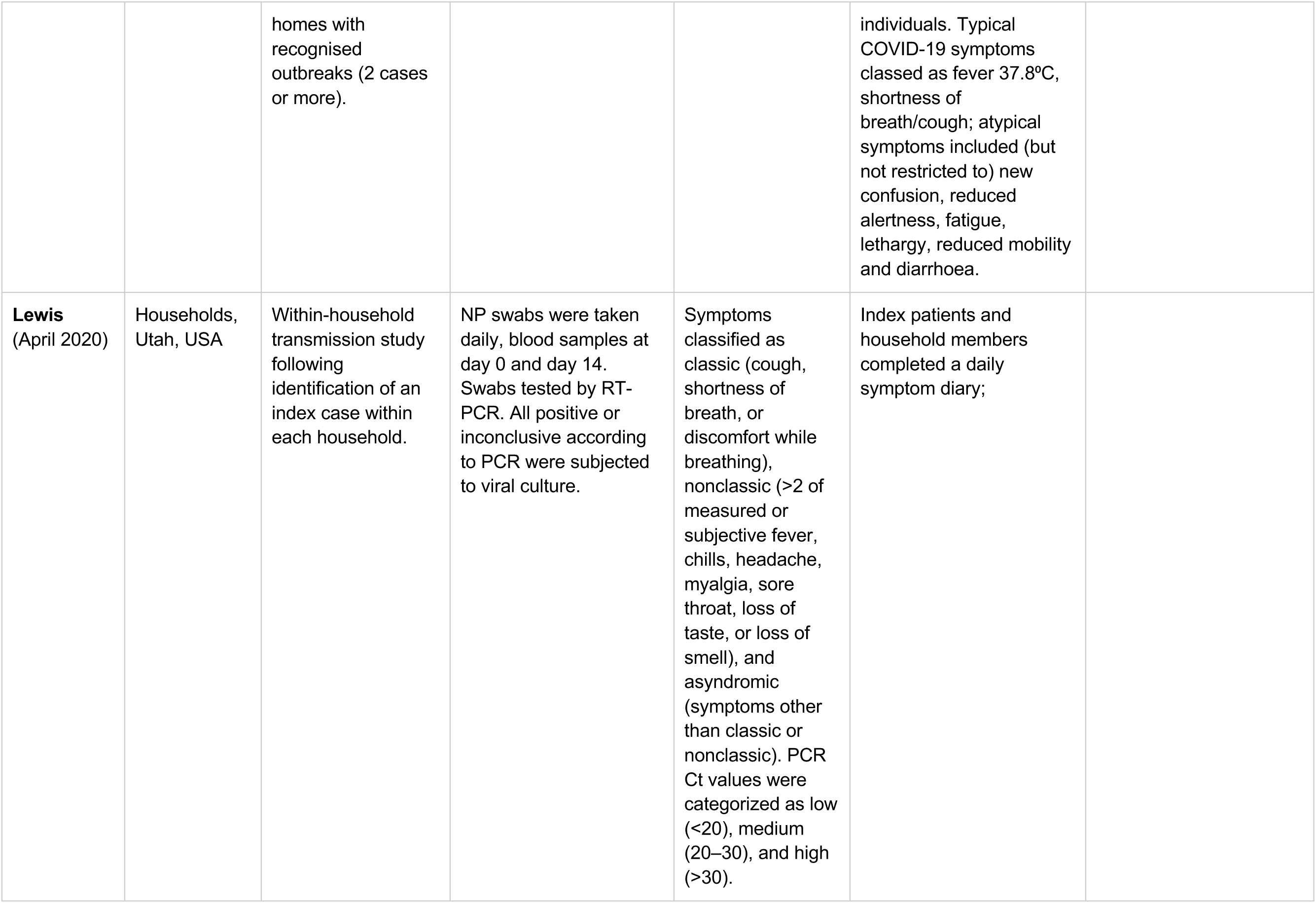

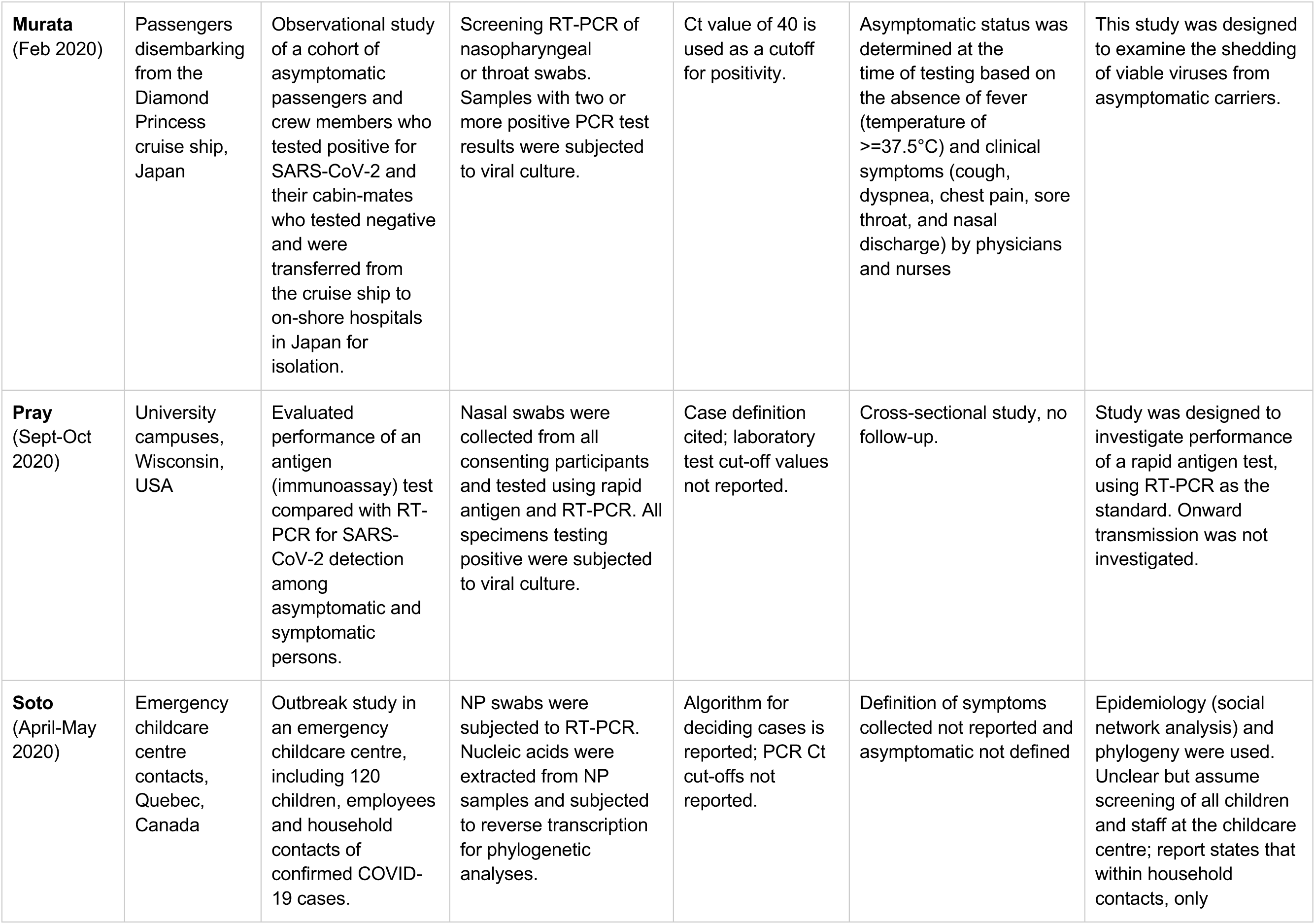

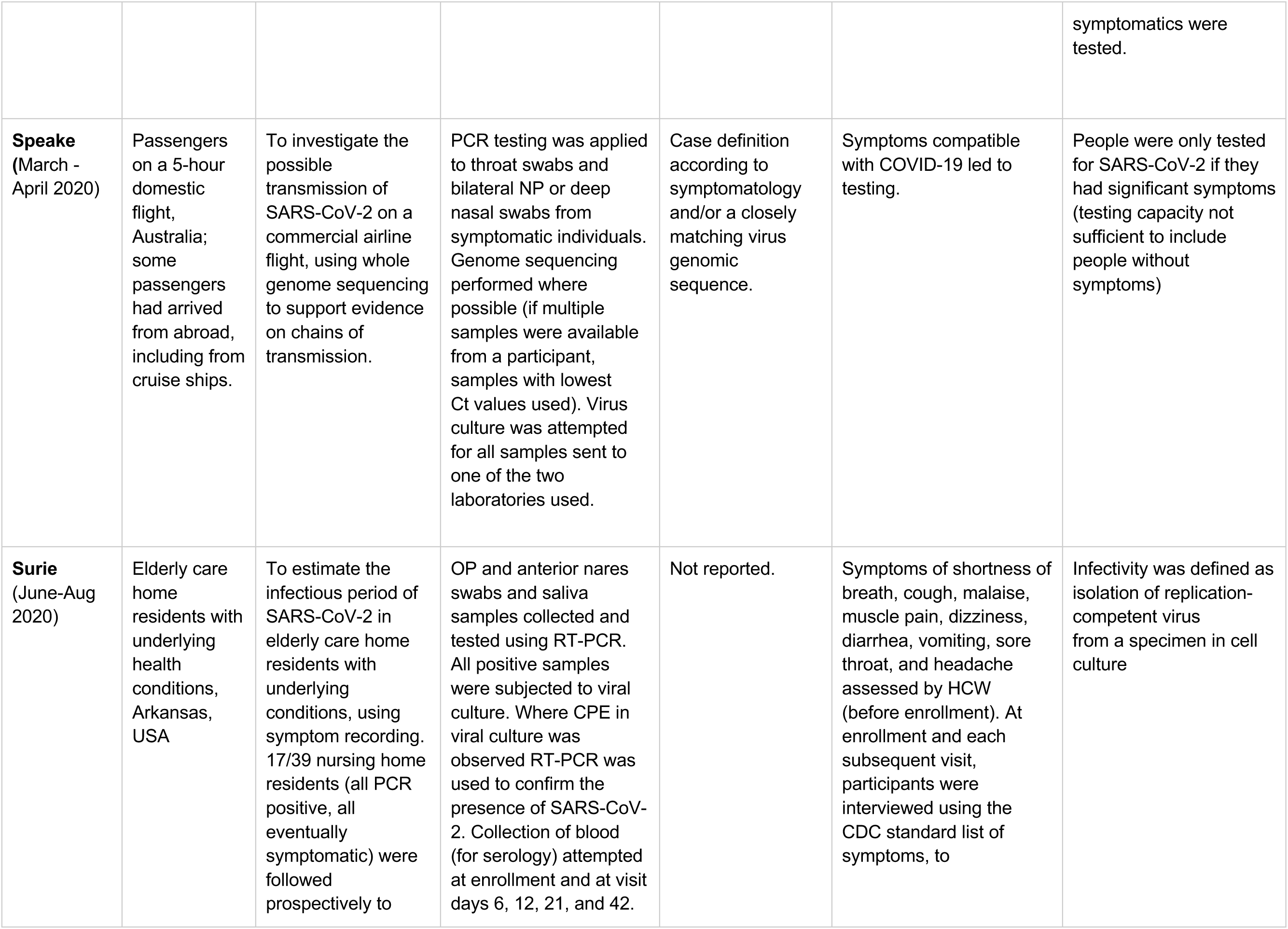

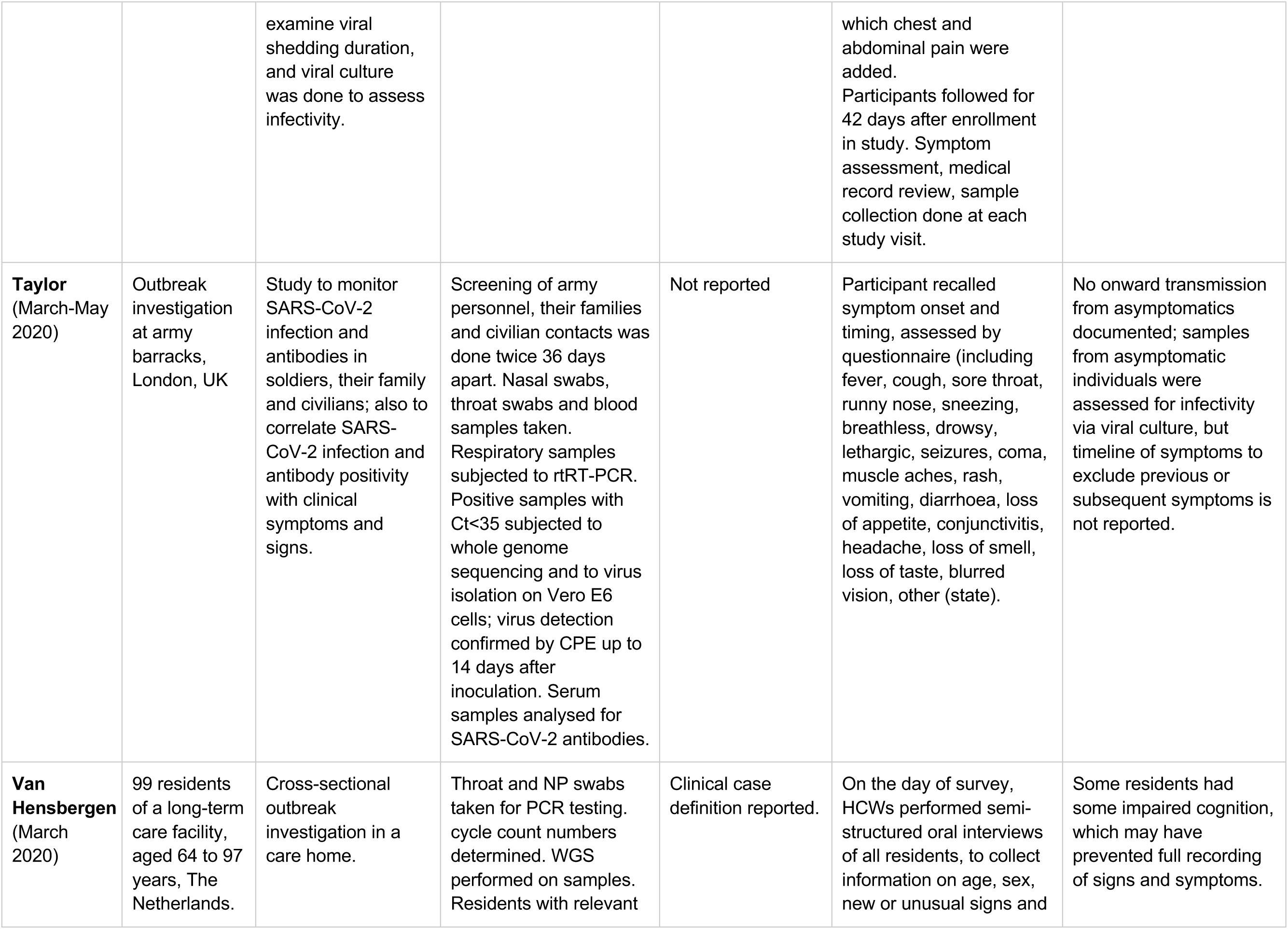

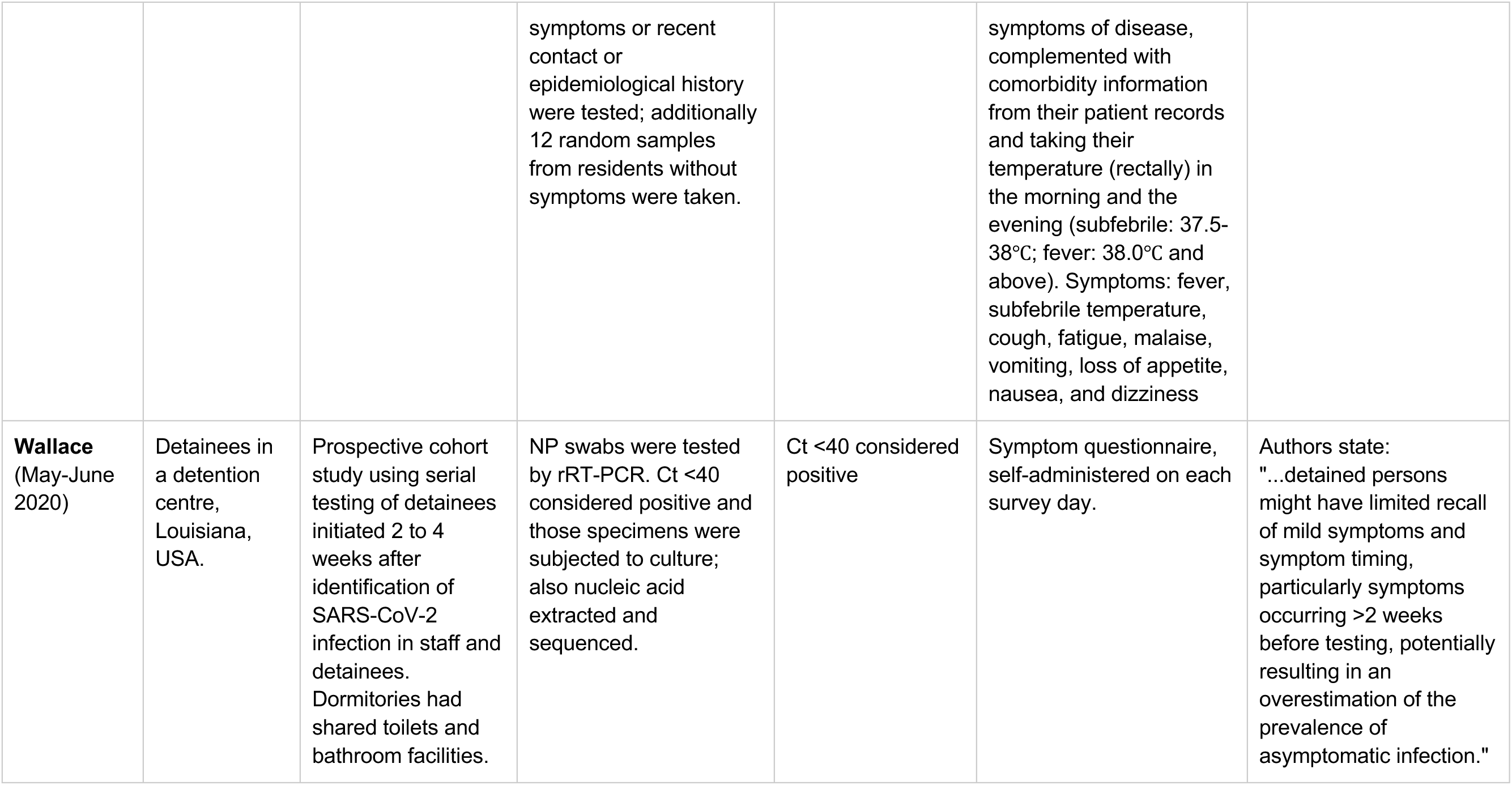

## Appendix C Quality Assessment of Symptom ascertainment

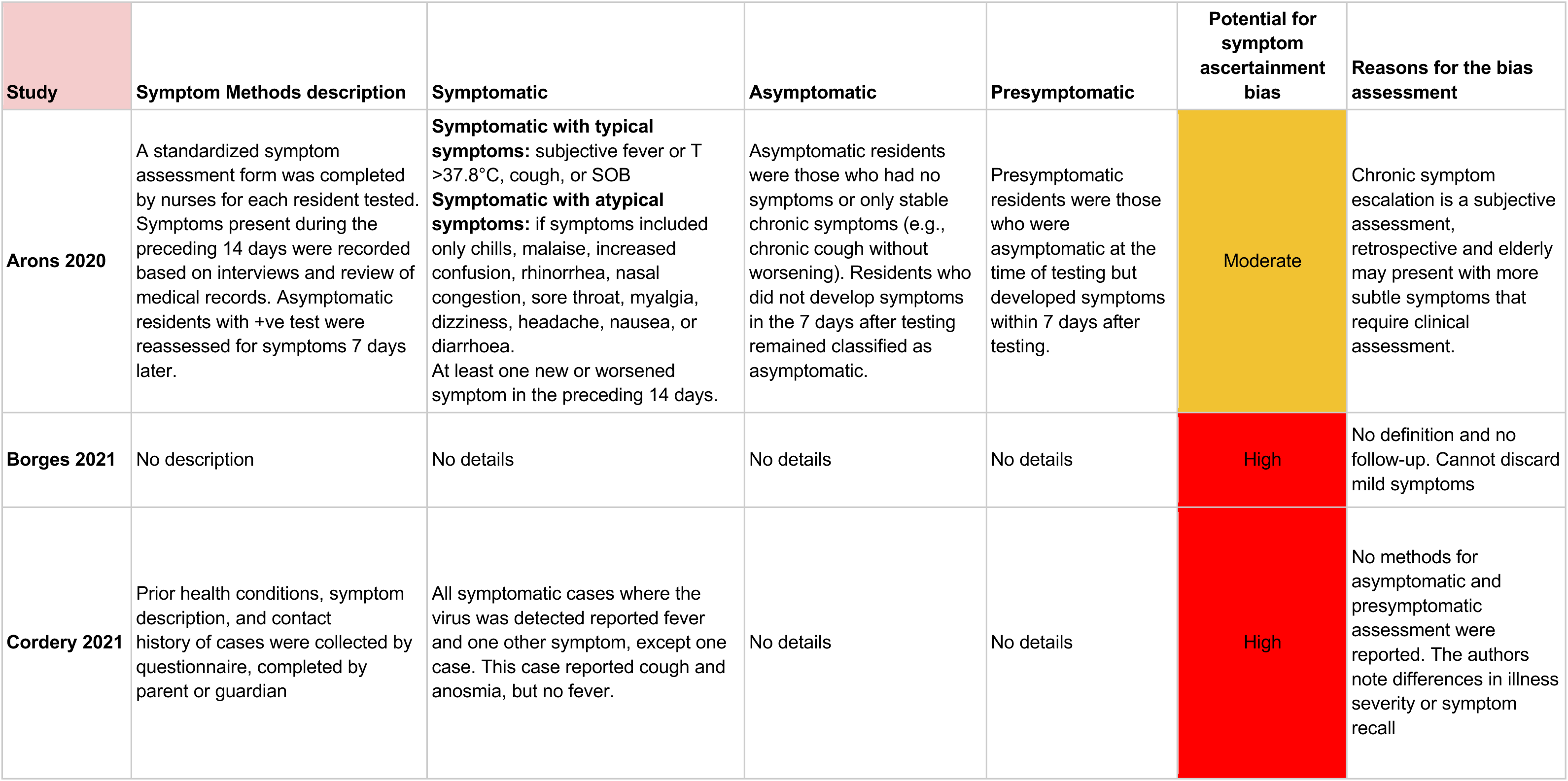

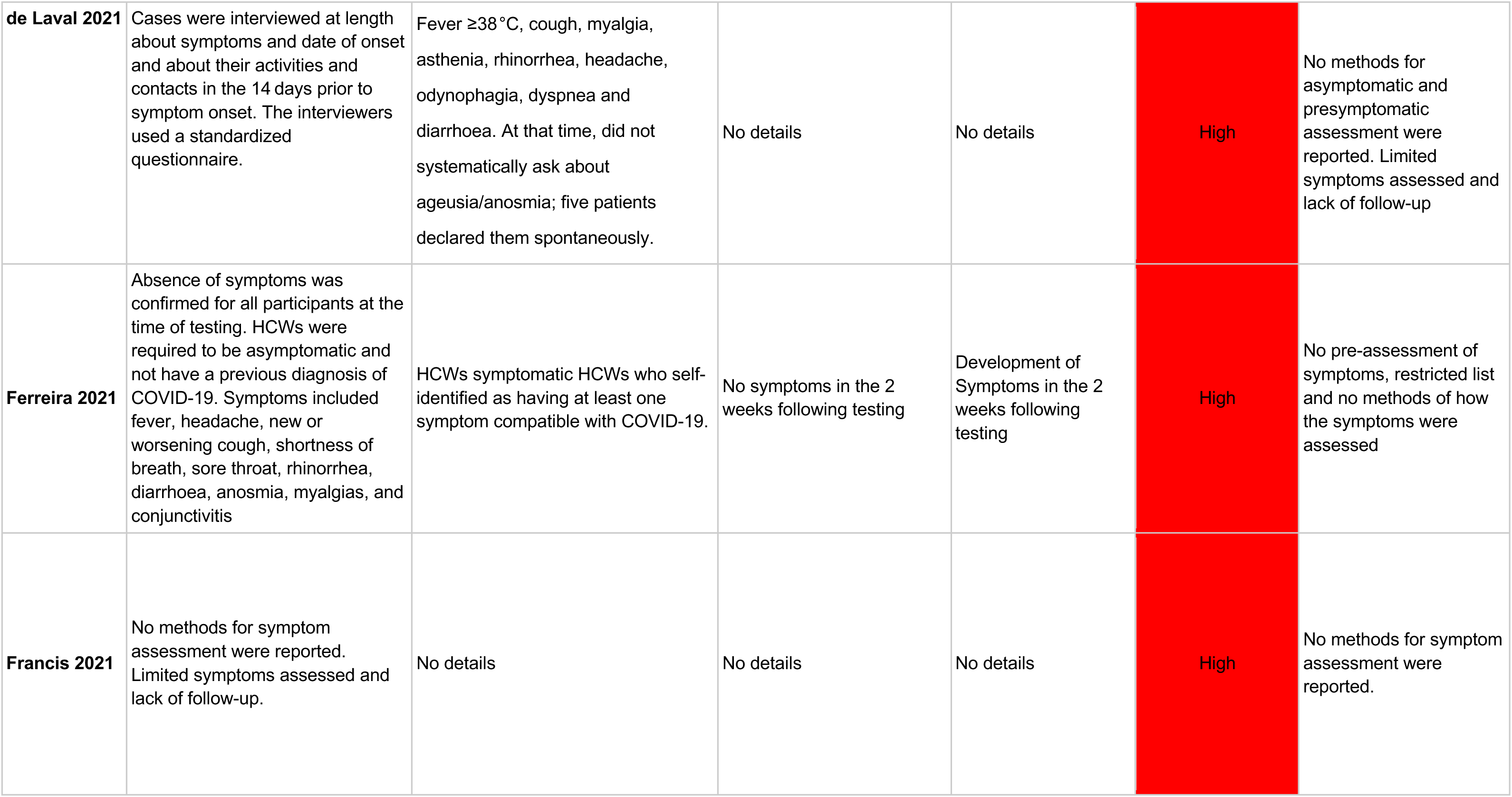

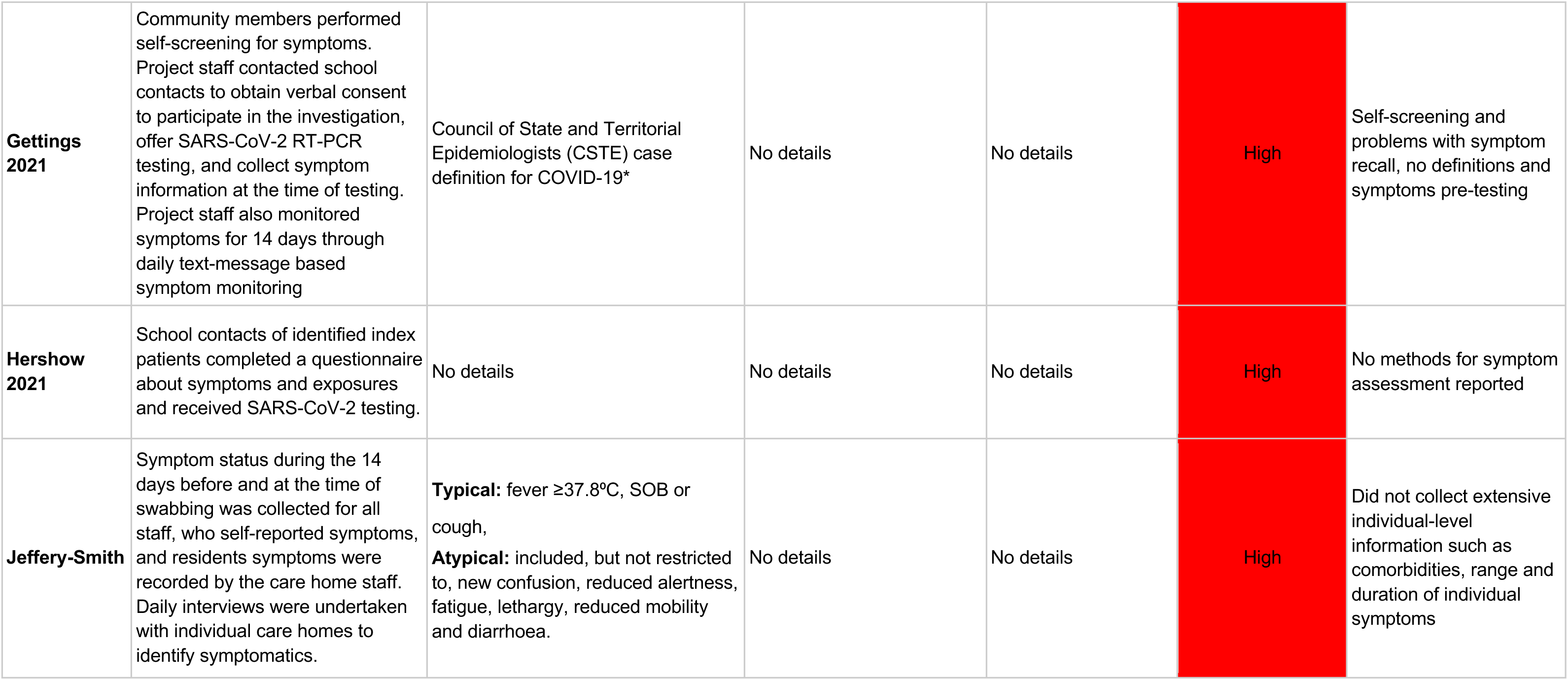

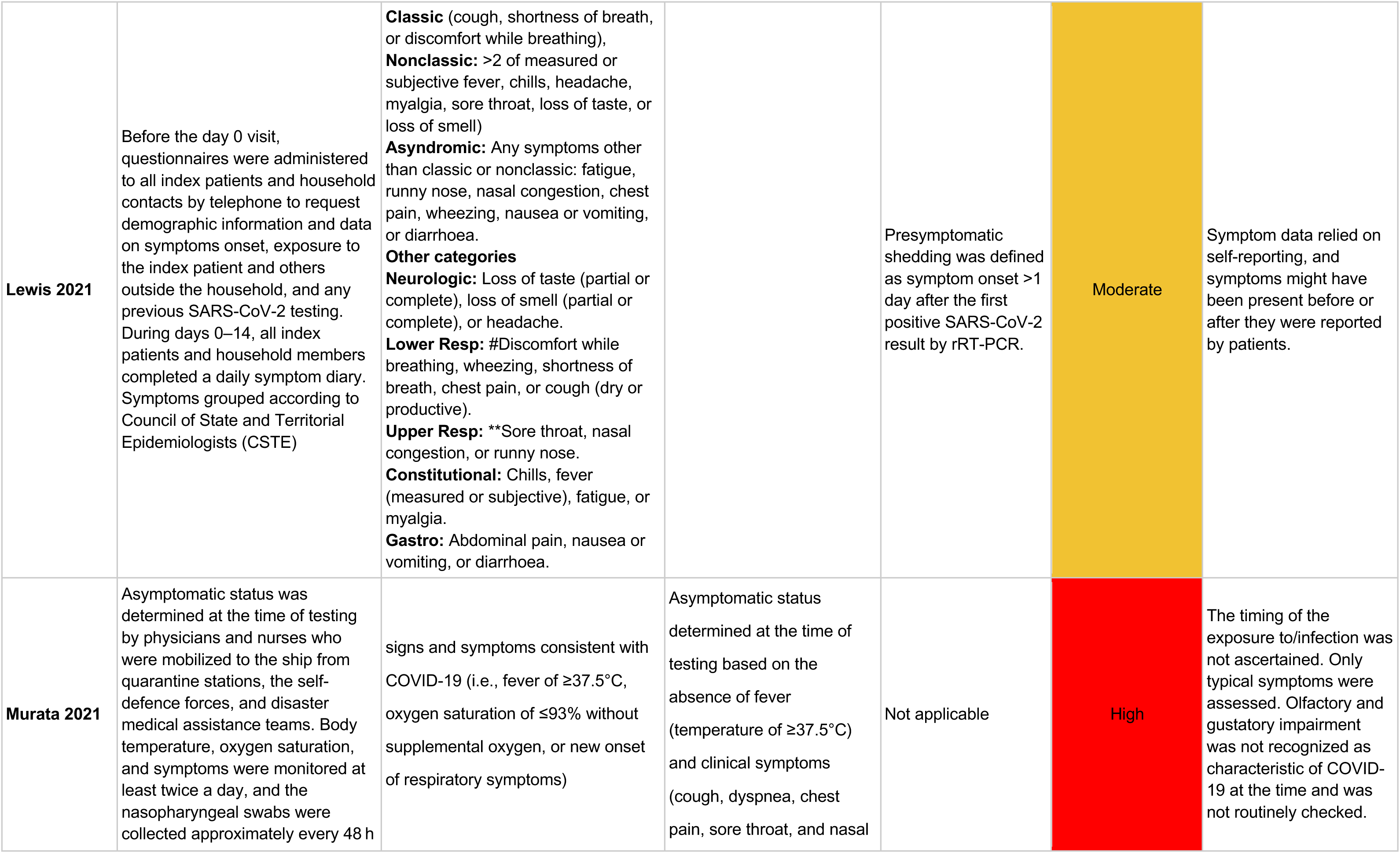

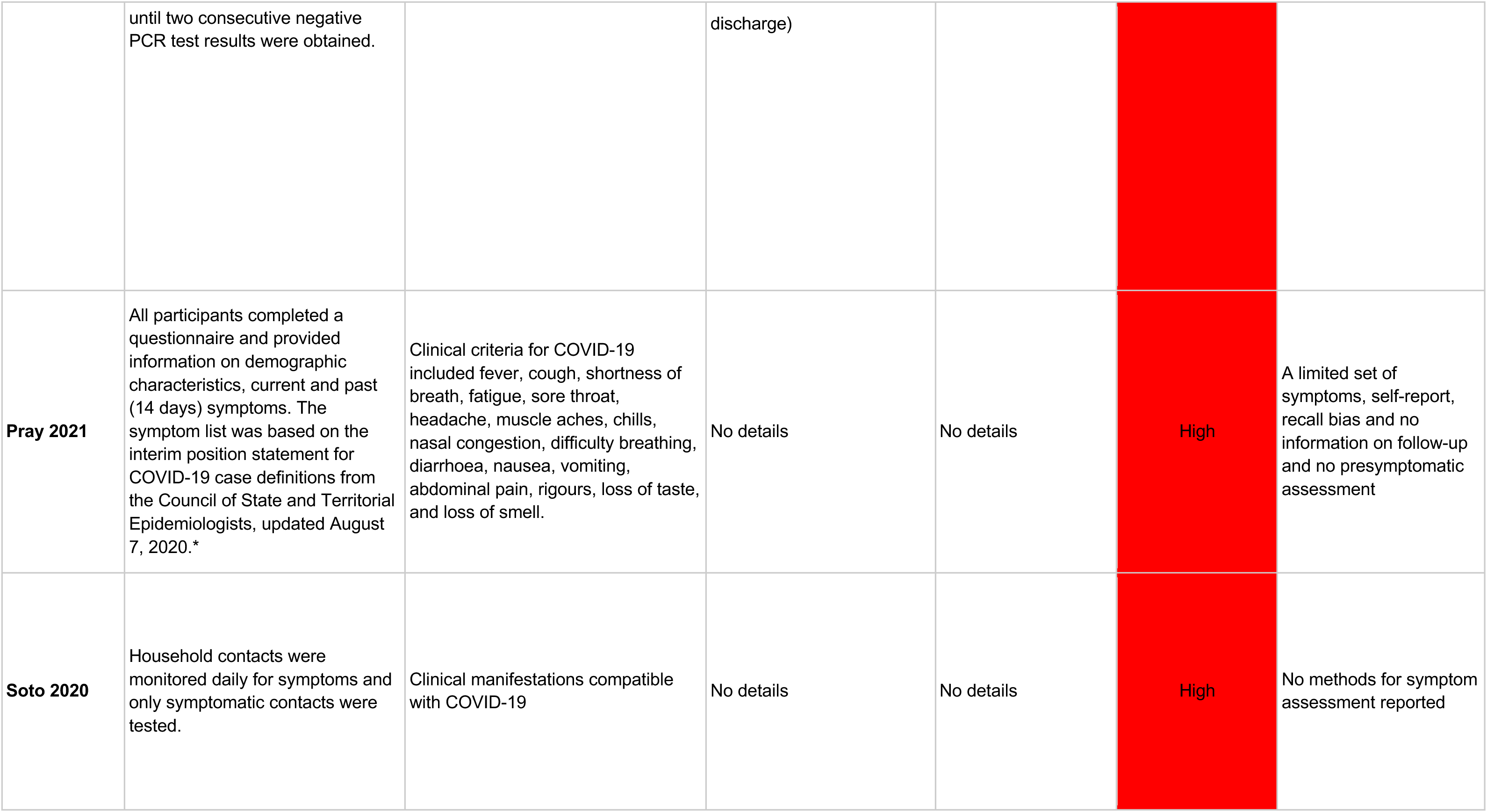

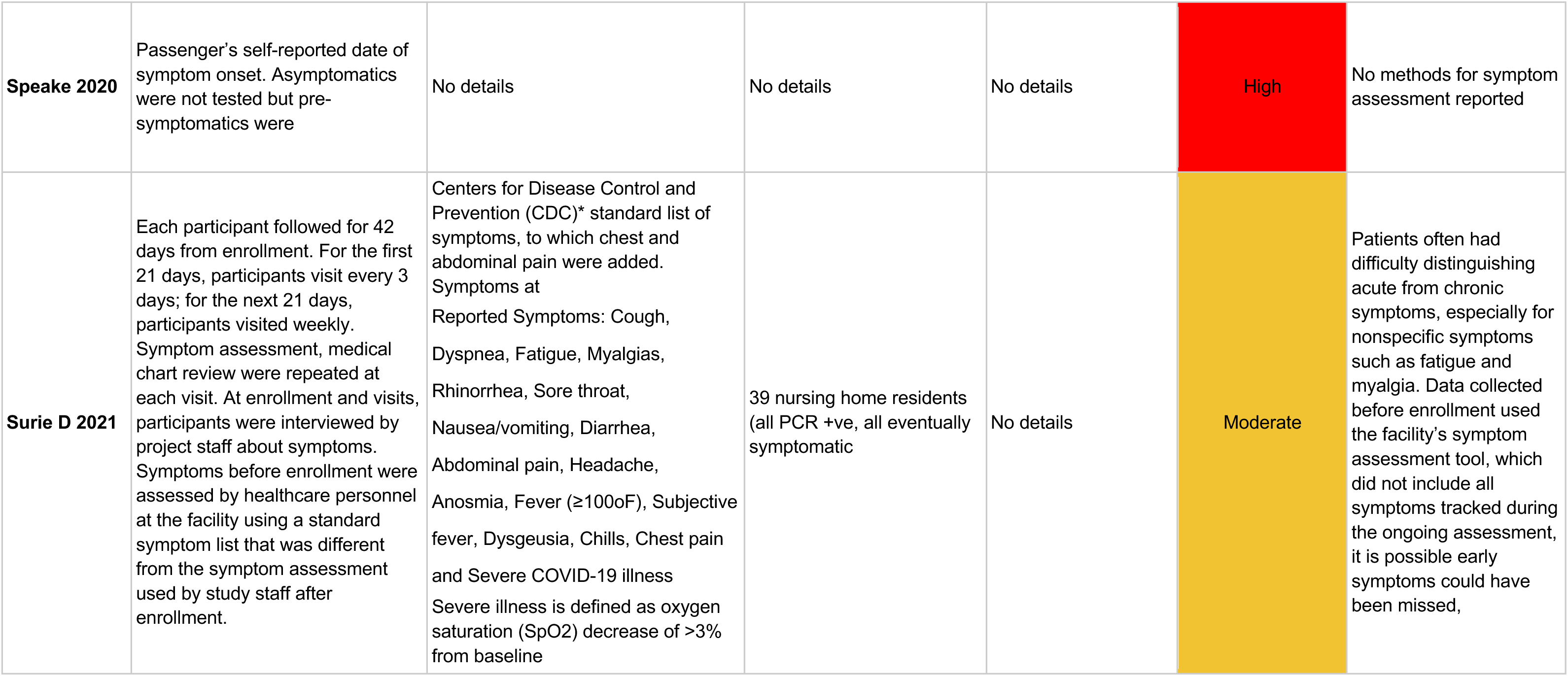

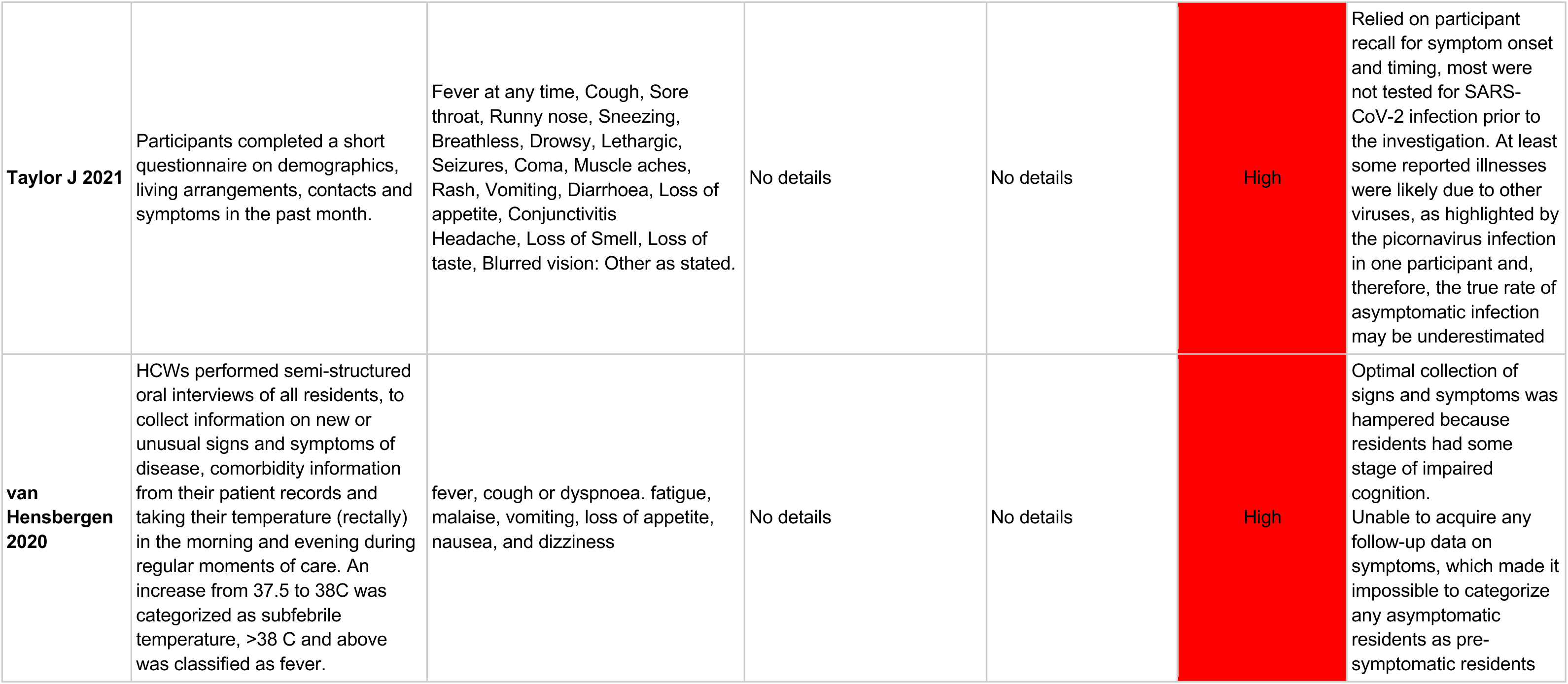

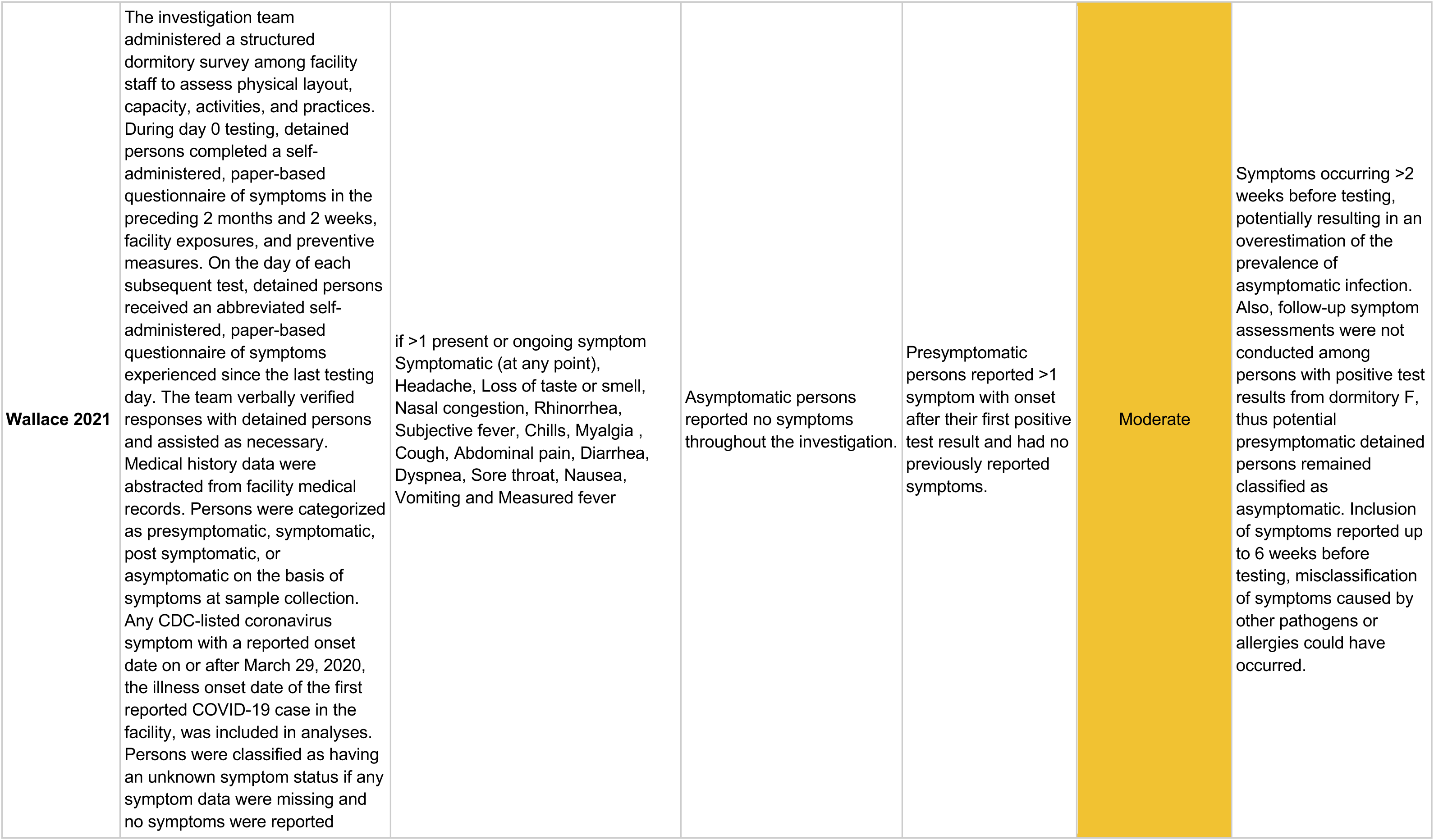

